# Obesity and Race Alter Gene Expression in Skin

**DOI:** 10.1101/2020.06.02.20120469

**Authors:** Jeanne M. Walker, Sandra Garcet, Jose O. Aleman, Christopher E. Mason, David Danko, Simone Zuffa, Jonathan R. Swann, James Krueger, Jan L. Breslow, Peter R. Holt

**Author notes:** These authors contributed equally to this work (Jeanne M. Walker, Sandra Garcet).

## Abstract

Obesity is accompanied by dysfunction of many organs, but effects on the skin have received little attention. We studied differences in epithelial thickness by histology and gene expression by Affymetrix gene arrays and PCR in the skin of 10 obese (BMI 35-50) and 10 normal weight (BMI 18.5-26.9) postmenopausal women paired by age and race. Epidermal thickness did not differ with obesity but the expression of genes encoding proteins associated with skin blood supply and wound healing were altered. In the obese, many gene expression pathways were broadly downregulated and subdermal fat showed pronounced inflammation. There were no changes in skin microbiota or metabolites. African American subjects differed from Caucasians with a trend to increased epidermal thickening. In obese African Americans, compared to obese Caucasians, we observed altered gene expression that may explain known differences in water content and stress response. African Americans showed markedly lower expression of the gene encoding the cystic fibrosis transmembrane regulator characteristic of the disease cystic fibrosis. The results from this preliminary study may explain the functional changes found in the skin of obese subjects and African Americans.

## Introduction

Obesity, defined as a body mass index (BMI) greater than 30 kg/m^2^ [1], has become a major epidemic in industrial and emerging countries. The prevalence of obesity has doubled since the 1980’s and it is now estimated that 600 million adults worldwide are obese [2]. Obesity affects many organs of the body and it is this organ dysfunction that leads to excess mortality and morbidity [3]. Much attention has focused on the consequences of obesity in the heart, liver and pancreas and other organs in which increased inflammation and oncogenesis become apparent [4]. Less attention has been paid to the effects of obesity on the skin.

Obesity increases psoriasis [5], which can be ameliorated with weight loss, and cutaneous infections [6]. Since diabetes is common in obesity, disorders such as fibroepithelial polyps and acanthosis nigricans also occur in the skin of obese subjects [7, 8]. Moreover, physiologic changes found in obese skin include increased trans-epidermal water loss with lower capacitance, dry, rough textured skin with pronounced erythema and reduced microvascular reactivity. Altered collagen formation and increased delayed-type hypersensitivity have also been reported [9].

Adipocyte depots that exist adjacent to the epidermis have distinct morphology and physiologic characteristics and are termed dermal or subdermal adipose tissue. In addition to the principal role for dermal adipocytes in lipid storage and thermal insulation [10], they also promote skin immunity [11], wound healing, and hair follicle cycling [12]. Obesity is accompanied by inflammatory immune changes in subcutaneous and visceral adipose tissues [13], but the role of inflammatory changes within the adipose layer of the skin has received little attention. Furthermore, obesity is associated with increased circulating leptin levels which appear to independently affect dermal cell proliferation and hair growth [14]. In addition, the microorganisms that live on the skin surface also affect skin immunity [11] so that it is important to analyse the skin microbiome comparing obese and normal individuals.

In view of the profound clinical and physiologic changes described in the skin in obesity, it would not be surprising also to find biologically important molecular changes. The present study was designed to compare gene expression in skin of healthy obese and non-obese subjects and to evaluate the potential importance of parallel changes in the microbiome and metabolites found on the adjacent skin surface and in the adipose tissue immediately below the skin.

## Materials and Methods

### Subjects

Participants were recruited from the surrounding community through advertisements and from the Rockefeller University subject repository. Eligible were healthy, obese (BMI 35-50 kg/m^2^) and non-obese (BMI 18-27 kg/m^2^) postmenopausal women between the ages of 40 and 70 years. The two groups were matched by age (+/- 5 years) and defined by self-reported race and by skin colour. Exclusions were unstable weight (> 5% change within the past three months), HIV infection, weight loss surgery, inflammatory bowel disease, history of malignancy other than non-melanoma skin cancer in the previous 5 years, generalizable or systemic skin diseases, history of a bleeding disorder, current anticoagulant therapy or regular NSAID use, current weight control medication or hypoglycaemic therapy, individuals taking oestrogen/progesterone hormones and current tobacco smokers. Also excluded were candidates with fasting blood glucose >125mg/dl, liver function tests (ALT, AST, alkaline phosphatase) greater than 2 times the upper limit of normal (ULN), abnormal thyroid function test, or serum creatinine > 2x ULN.

Fourteen obese subjects were screened, two refused skin biopsies, one was withdrawn due to an intercurrent inflammatory illness, and one was not postmenopausal by our criteria. Ten obese subjects met our inclusion criteria and underwent skin swab collections and punch biopsy. Twenty non-obese subjects were screened. Two subjects refused to undergo punch biopsy, one was withdrawn due to an intercurrent illness, two with a BMI outside the required range, one withdrew consent, one was excluded with a history of keloid formation, one with a low platelet count, one with uncontrolled hypertension. One non-obese subject who underwent skin punch biopsy was not included in the analysis because we were unable to find an age and race-matched obese subject. These 10 obese and 10 age-matched, race-matched non-obese postmenopausal women completed all aspects of the study (Figure 1). Six participants were Caucasian and four were African Americans in each group. Post-menopausal women were chosen to exclude effects of the menstrual cycle upon study end points and to exclude gender effects.

**Figure 1.**
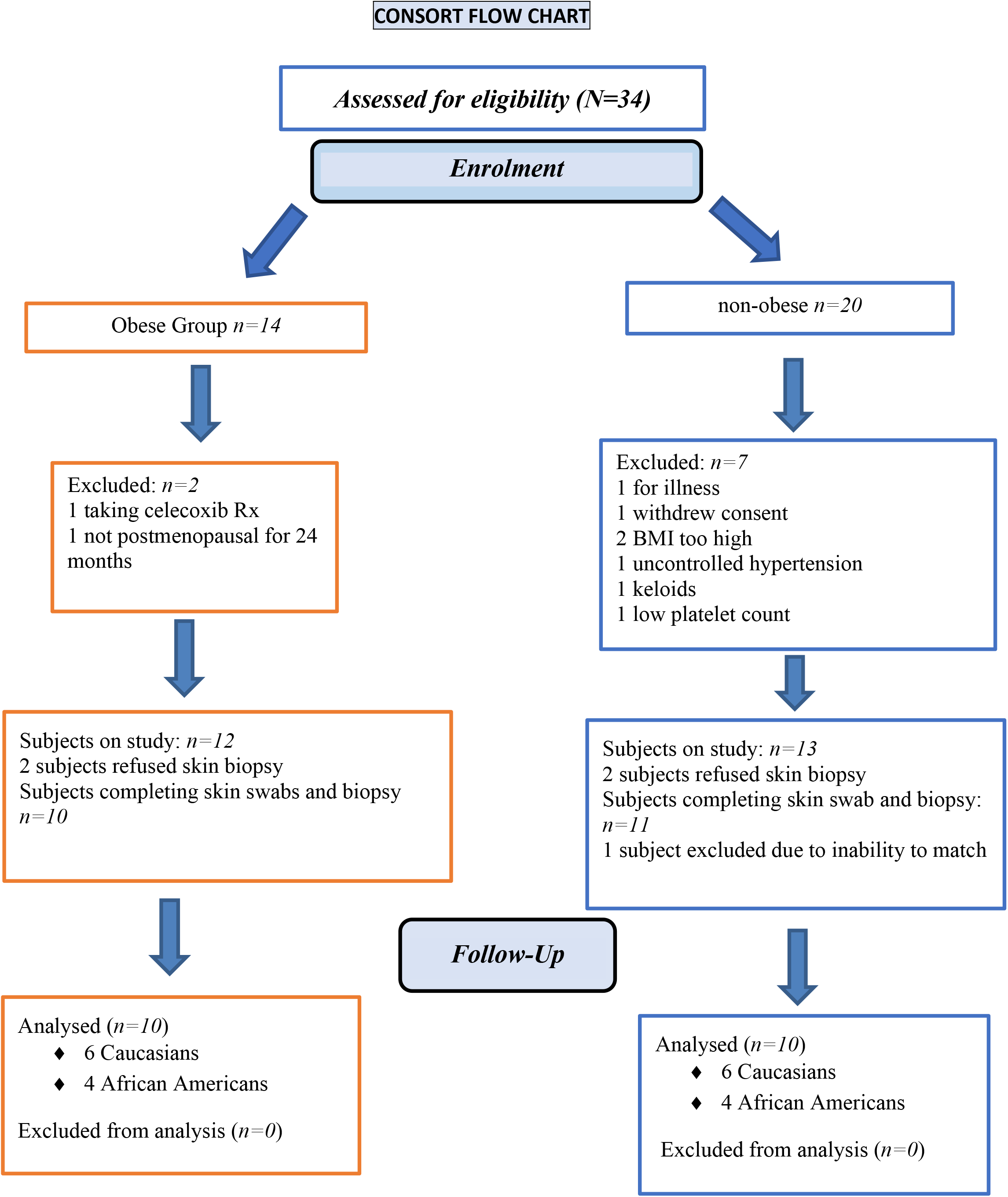
Consort flow chart of eligible subjects

Based on preliminary data from a previous study comparing skin from seven obese and six non-obese postmenopausal women, there was a variation of 45% in a set of RT-PCR genes (unpublished data). Assuming the same variation and proportion of differentially expressed genes to be 5%, we calculated that a sample size of *n* = 10 subjects per group, matched by age and race, would provide 80% power at a 5% false-discovery rate to detect the expected number of differentially expressed genes based on a threshold of 2-fold changes.

### Design and Setting

This was an open-label comparison of a group of 10 postmenopausal obese women and 10 postmenopausal non-obese women who were age-matched (+/- 5 years) and race-matched. Screening comprised a complete history and physical examination and fasting blood testing for complete blood count, sedimentation rate, comprehensive chemistry panel, lipid panel, thyroid function tests, hepatitis C antibody, uric acid, and haemoglobin A1C. Observing Good Clinical Practice guidelines, all participants read and signed an informed consent document approved by the Institutional Review Board and the Advisory Committee for Clinical and Translational Science at The Rockefeller University (Protocol JWA-0921).

## Procedure Methods

### Anthropometric measurements

Body weight was measured daily to the nearest 0.1 kg using a Scale-Tronix 5002 scale (Welch Allyn, Skaneateles Falls, N.Y.) with precision of +/- 0.1kg. Subjects were weighed in a hospital gown, after an overnight fast and post-voiding. Height was measured to the nearest 0.1 cm at baseline with a Seca-216 stadiometer (Hamburg, Germany) in 1 cm increments. Body mass index (BMI) was calculated as (kg/m^2^), using the NIH Standard Metric BMI calculator.

### Blood collection and analysis

Fasting blood samples were analysed in the Clinical Pathology Laboratory of the Memorial Sloan-Kettering Cancer Center for complete blood count, electrolytes, glucose, creatinine, blood urea nitrogen, liver function, C-reactive protein, sedimentation rate, and uric acid. Research serum samples were drawn pre and post intervention, aliquoted and stored at −80° C for subsequent analysis.

### Skin swabbing

Subjects were permitted to shower but did not wash the planned biopsy area over the mid-lower abdomen with soap for 3 days before the biopsy.

For microbiome analysis, two areas of skin approximately 10 × 10 cm were swabbed using the eSwab collection and preservation system for aerobic, anaerobic, and fastidious bacteria (Copan Diagnostics, Marietta, CA). Swabs were labelled, sealed separately in the provided tubes, and immediately stored at −80° C. For metabolome analysis, two different areas of skin approximately 15 × 15 cm were swabbed using saline-moistened sterile cotton-tipped applicators. The tips were cut, sealed in separate sterile collection tubes, and immediately stored at −80° C. Details of the analyses of the microbiome and metabolome skin swabs are found in the supplement under “Methods”. The scientists making these measurements and statistical analyses were blinded to the group assignments.

### Skin biopsy

After the skin swabbing, the abdominal site was cleansed with (3) Chloraprep swabs (chlorhexidine 2% and isopropyl alcohol 70%, Becton Dickinson, Canaan, CT). Using sterile technique, local anaesthesia was induced by infiltration of the area with 4 ml of lidocaine 1 % (Hospira, Inc., Lake Forest, IL) mixed with 1 ml sodium bicarbonate. The skin biopsy was performed using a 6 mm punch (Miltex Instruments, York, PA). Fat tissue was carefully removed from the skin core of the biopsy using a scalpel. The skin core and fat were placed separately in RNAlater Stabilization Solution (Thermo Fisher Scientific, Fair Lawn, N.J.), refrigerated for 24 hours, and then frozen at −80° C. The biopsy site was sutured closed and a dry sterile dressing was applied. Subjects were discharged with wound care instructions and scheduled to return for suture removal.

### Gene-array and quantitative real-time PCR analysis

RNA was extracted, followed by hybridization to Affymetrix Human U133 Plus 2.0 gene arrays (Santa Clara, CA) or quantitative RT-PCR as previously described [15,16].

All statistical analyses were carried out in R (Limna) Log 2-transformed qRT-PCR measurements (hARP normalized) and microarray expression values were assessed with a mixed-effect. The fixed factors were condition (obese vs non-obese), race (African American vs Caucasian), with random intercept for each subject. Quality control of microarray chips was carried out using standard QC metrics and R package microarray quality control. Images were scrutinized for spatial artefacts using Harshlight [17]. Expression measures were obtained using the GCRMA algorithm [18]. A batch effect corresponding to the hybridization date was detected by principal components analysis (PCA) and adjusted using the ComBat function from the SVA package. Probe sets with at least 3 samples with expression values > 3 were kept for further analysis. Fold changes for the comparisons of interest were estimated, and hypothesis testing was conducted with contrasts under the general framework for linear models with the limma package. P values from the moderated (paired) t-tests were adjusted for multiple hypotheses using the Benjamini-Hochberg procedure. Hierarchical clustering was performed with Euclidean distance and a McQuitty agglomeration scheme [18, 19].

Data deposited into Gene Expression Omnibus (GEO) repository. GEO accession number pending.

## Results

This study was performed in ten healthy obese and ten healthy non-obese post-menopausal women matched for age and race. Obese subjects had a mean weight of 109 kg, BMI of 40.7 kg/m^2^, and waist circumference of 117 cm. Non-obese subjects had a mean weight of 59.4 kg., BMI of 22.4 kg/m^2^, and waist circumference of 80.4 cm (Table S1). The skin thickness for subjects with obesity was not significantly different from that of non-obese subjects (Figure 2).

**Figure 2.**
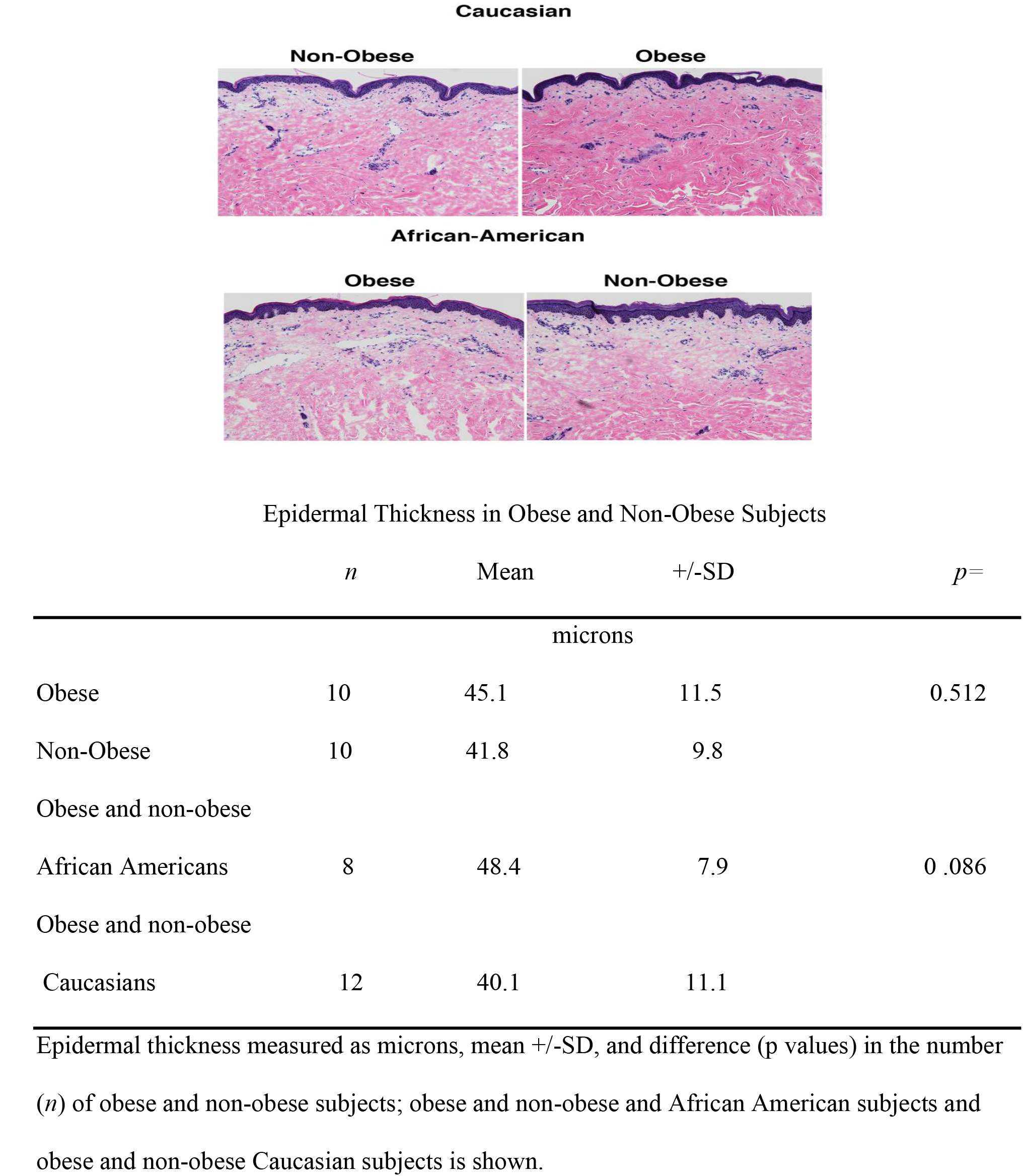
A. Typical photomicrograph of skin biopsies from obese and non-obese subjects and from African American and Caucasian subjects Paired by age and skin colour B. Dimensions of the epidermis of obese and non-obese African American and Caucasian subjects. Note no difference in epidermal thickness between obese and non-obese subjects but a trend to a wider epidermis in African Americans than in Caucasian subjects.

### Gene expression analysis of the skin

The 50 most differentially expressed genes in the skin between obese and non-obese subjects are displayed in the heat map in Figure 3A. Comparing gene expression in obese versus non-obese skin showed greater gene expression of S100A7A, encoding a calcium binding protein involved in psoriasis, and CORIN encoding a natriuretic peptide converting enzyme, which is expressed in the dermis and is involved in specifying skin colour. However, the expression of CREB3LA, encoding a cyclic AMP response element, was lower in the skin of obese subjects.

**Figure 3.**
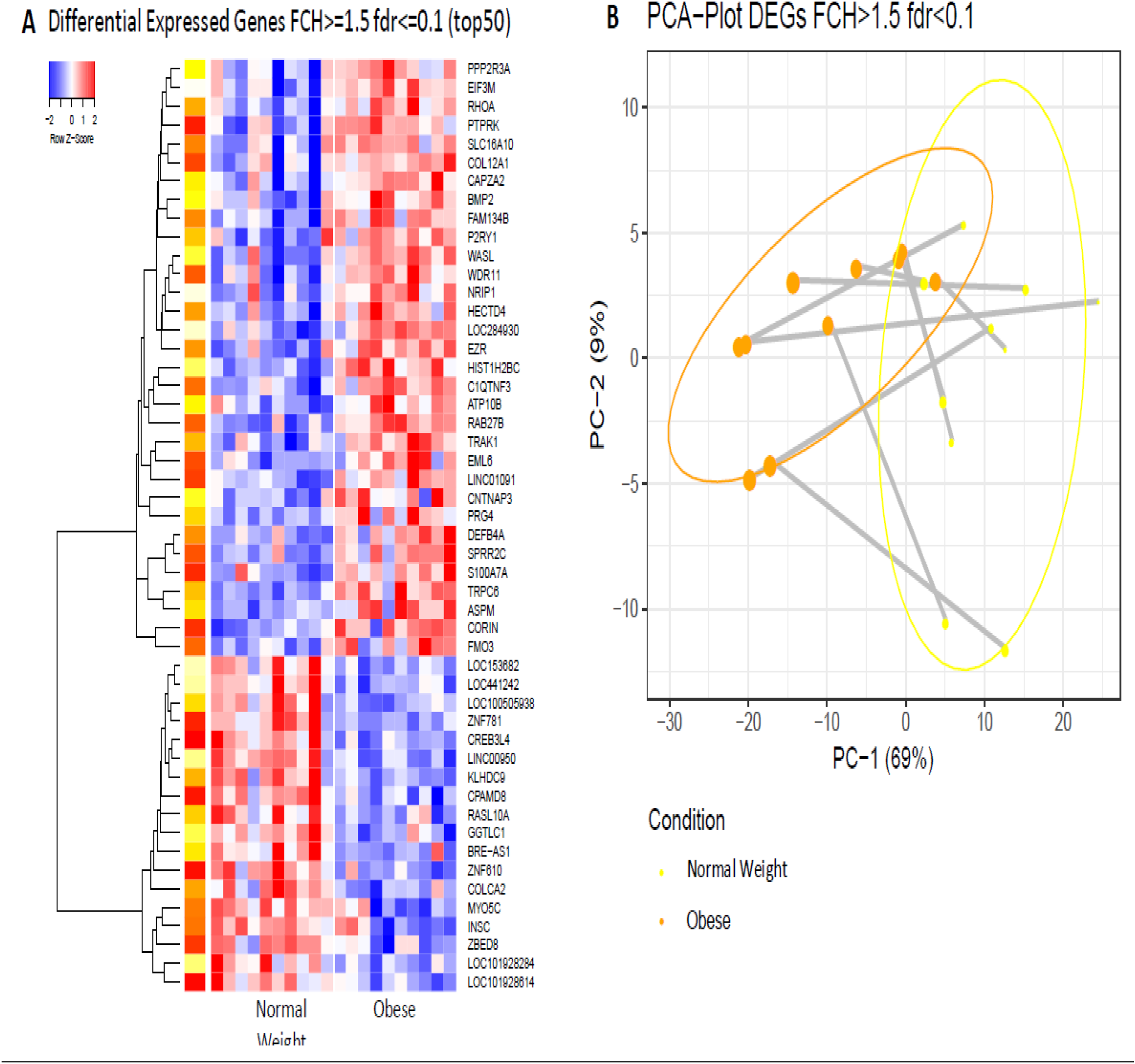
Differences in gene expression between the skin of obese and non-obese subjects A. Heat map of the 50 most differentially expressed genes in the skin of obese and non-obese subjects with an FCH (fold change)>=1.551; fdr (false discovery rate)<=0.1 B. PCA (principal component analysis) plot of differentially expressed genes in the skin of obese and non-obese subjects with an FCH>=1.5; fdr<0.1

A PCA model constructed on 400 of the most differentially expressed genes between obese and non-obese subjects showed partial separation of the 2 groups. This difference was seen in PC1 which accounted for 69% of the variation in the included genes (Figure 3B).

The complete list of skin genes whose expression significantly differed between the two groups is shown with fold changes in Table S2. Again, the gene expression of S100A7A was 3.44-fold higher in the obese skin compared to the non-obese skin. Similarly, the expression of DEFB4A (Defensin B4A), which encodes an antimicrobial peptide (part of the beta-defensive system), and SPRR2C, which encodes a proline rich protein strongly induced during differentiation of human epidermal keratinocytes, was also significantly higher in the obese skin being 3.11 and 1.7fold higher, respectively. Genes with lower expression profiles in obese subjects than non-obese included AOP that encodes aquaporin (involved in water channels present in the skin), PROM1 (prominin 1; involved in cell differentiation and proliferation) and Keratin 7 and 19 (important for fibrogenesis in the epidermis). Also, of interest was the significantly higher expression (2.82-fold) of CFTR, the cystic fibrosis transmembrane conductance receptor in non-obese subjects compared to the obese group.

QTPCR analysis of 15 genes selected from the total list of significantly differentially expressed genes in skin confirmed increased expression of the S100A (3.73-fold), DEFB4A (defensin B4A) (3.29-fold), and CORIN (2.07-fold) in the skin of obese subjects (Figure 4). Significantly lower gene expression in the skin of obese subjects was found with CFTR, (3.6-fold), PROM1 (5.56-fold), and GABRP (gamma aminobutyric acid receptor) (2.9-fold).

**Figure 4.**
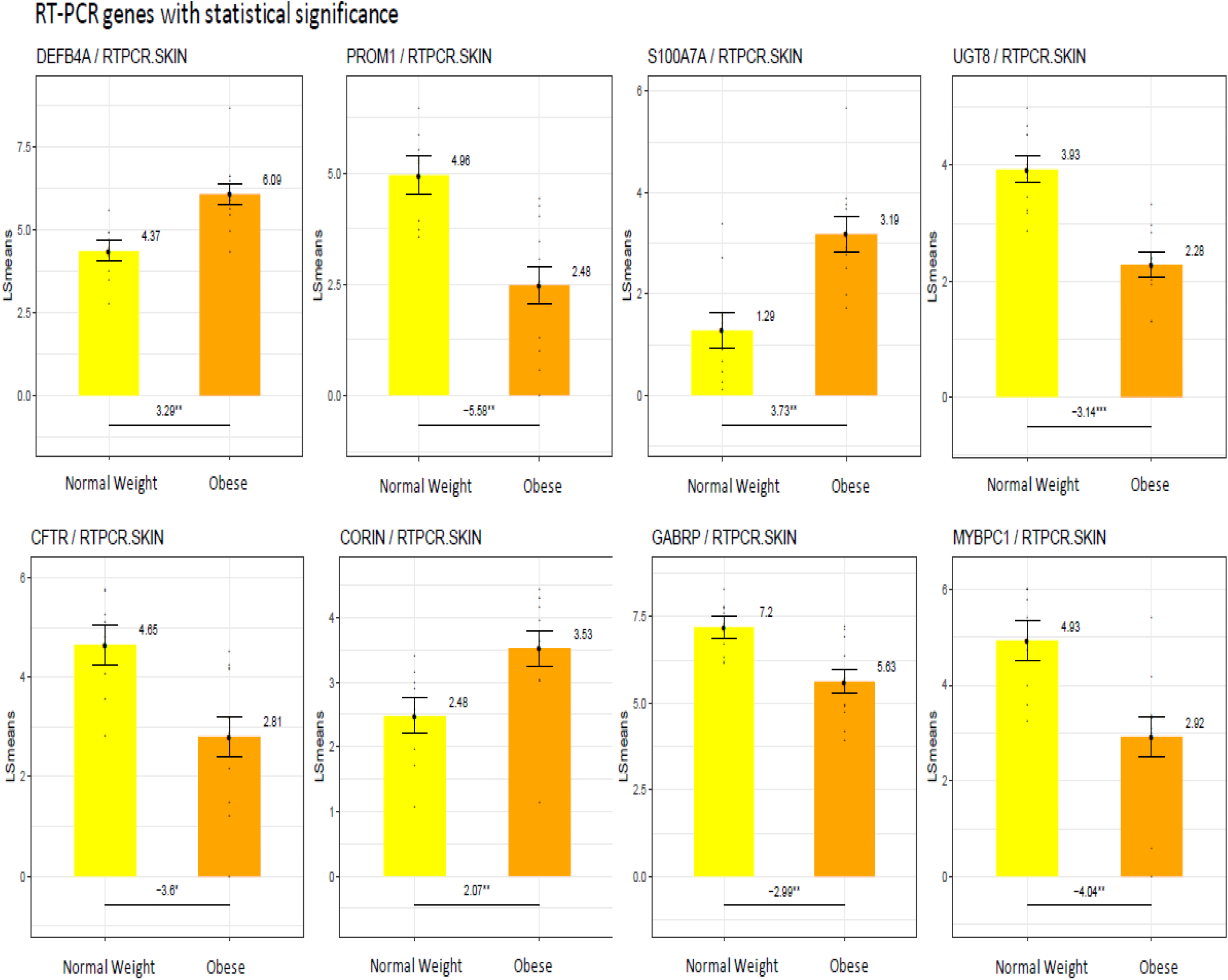
Differences in gene expression by RT-PCR between the skin of obese and non-obese subjects. LS means of gene expression by RT-PCR showing significant differences as *p<0.05; **p<0.01; ***p<0.001

Gene expression pathway analysis showed a broad downregulation of many pathways in obesity subjects with only 4 out of 39 of the most highly differentially expressed pathways higher in the obese (Figure 5). The pathways most down-regulated included cardiac beta-adrenergic signalling, which appears to function in skin, cyclin dependent Kinase 5 (CDK5), a mutation which is important in melanoma formation and functions in skin healing, and gonadotrophic releasing hormone (GNRH) signalling, which has many extra pituitary functions.

**Figure 5.**
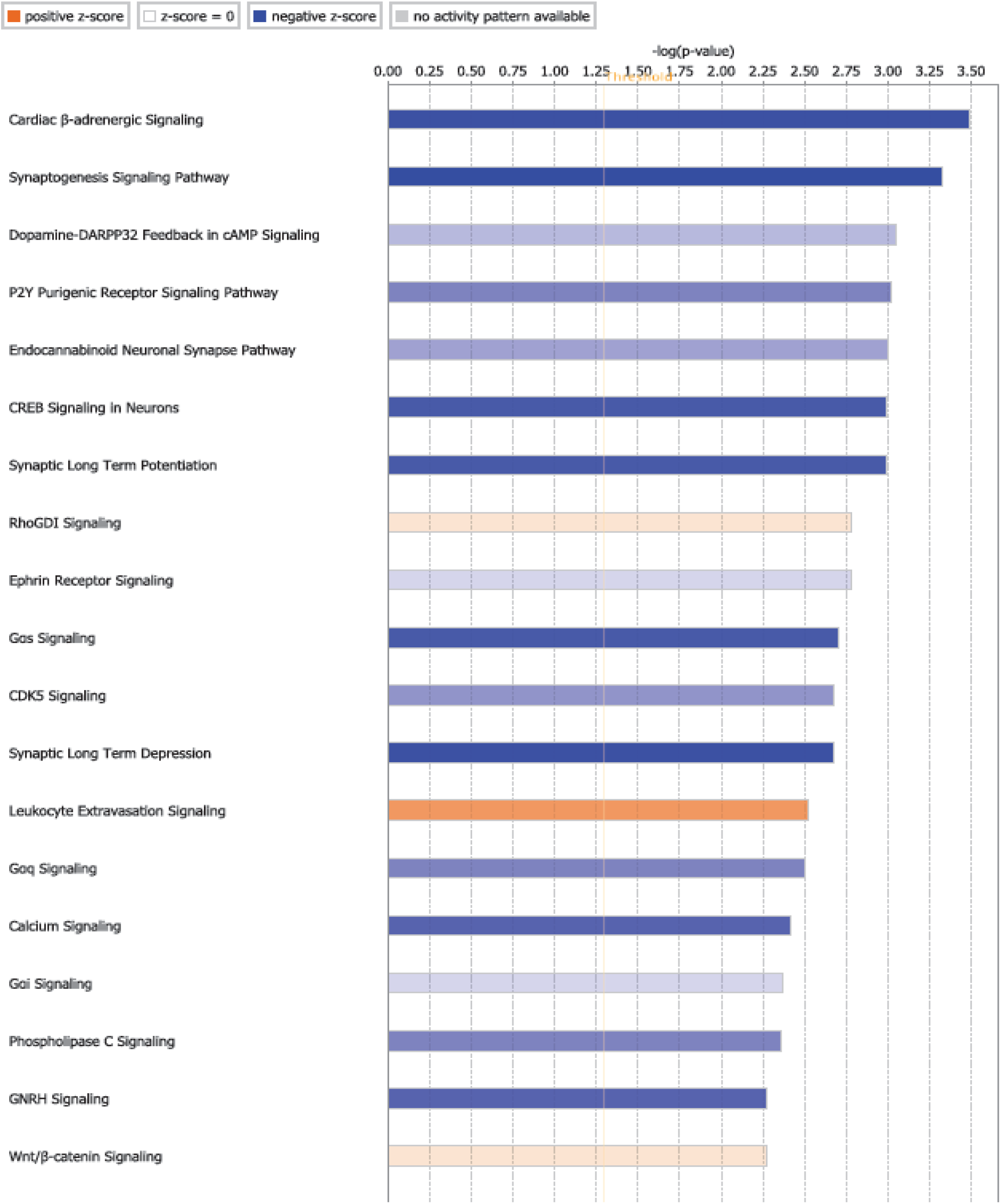
Gene expression pathway analysis between the skin of obese and non-obese subjects. Data shown from analysis of all differentially expressed genes. Log value shown in blue = negative *z* score Log value shown in orange = positive *z* score.

### Gene expression analysis in skin fat

We next examined differences in gene expression between the 2 groups of subjects in subdermal fat removed from immediately below the skin portion of the biopsy. A heat map of gene expression shows a markedly different pattern between the two groups (Figure 6A). The expression of many of the genes upregulated in obese subdermal fat are involved with inflammation and immune function including platelet activating factor PLA2G7, ILIRN involved in IL1 activation, SPPI, a cytokine that can increase interferon gamma and IL12 activity, and several serpins, mediators involved in inflammation and immune function.

**Figure 6.**
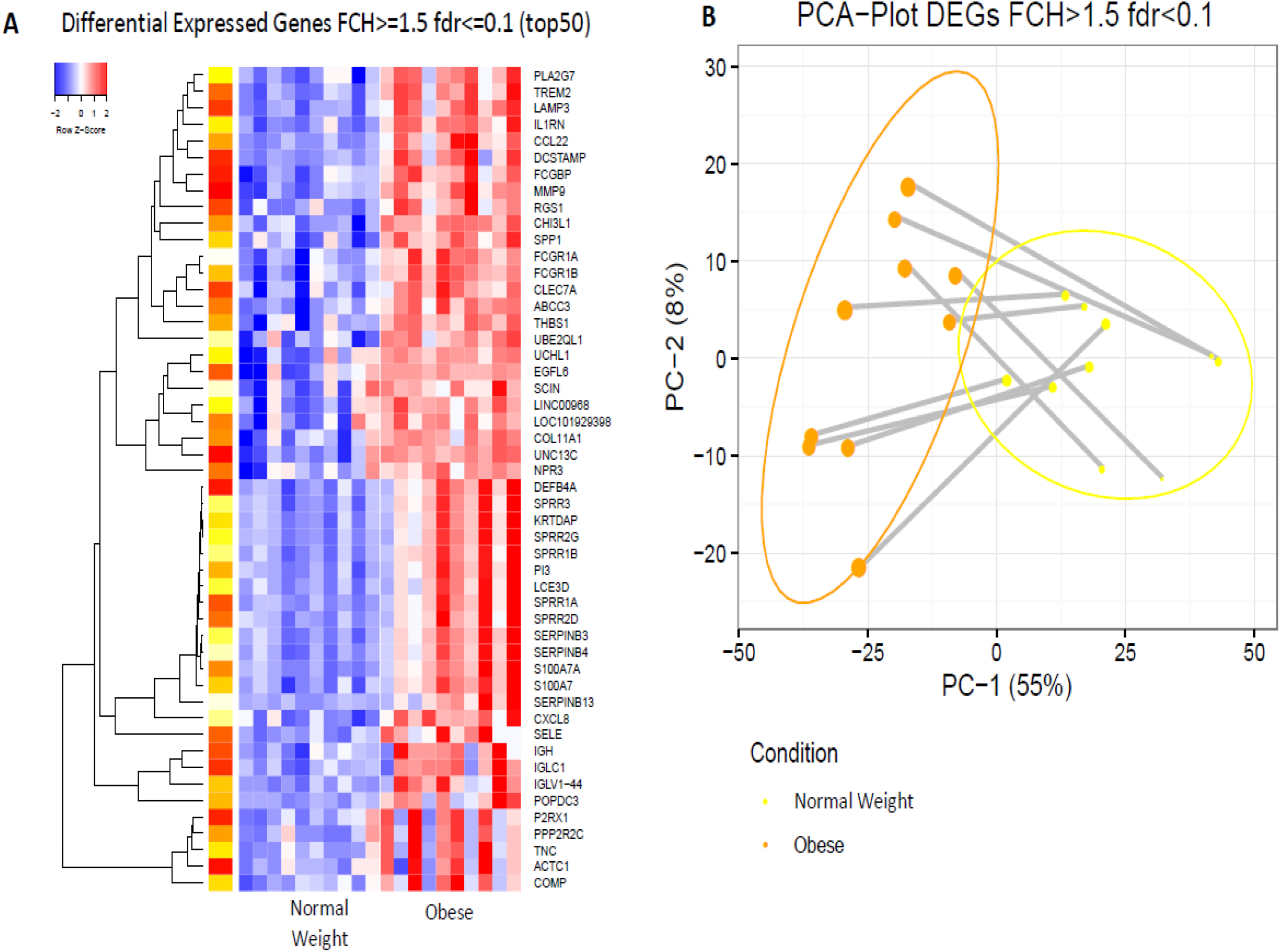
Differences in gene expression between the subdermal fat of obese and non-obese subjects A. Heat map of the 50 most differentially expressed genes in subdermal fat of obese and non-obese subjects with an FCH>=1.5; fdr<0.1 B. PCA plot of differentially expressed genes in subdermal fat of obese and non-obese subjects with an FCH>=1.5; fdr<0.1

The PCA plot of genes whose expression differed significantly in subdermal fat of obese and non-obese subjects (Figure 6B) clearly shows separation between the two groups. Most of the difference was seen in PC1 which includes 55% of the genes whose expression was determined.

Table S6 shows a list of genes whose expression was relatively greater in the subdermal fat of obese subjects. The expression of SPPI that encodes osteopontin, which can act as a cytokine, augmenting the action of interferon gamma and interleukin 12 was approximately 10-fold higher in the obese subjects. EGFL6 expression, which encodes an epidermal growth factor found to be enhanced in obesity and alters insulin action, was increased by 8.5-fold. MMP9, which encodes metalloproteinase 9 and ILTRN, was increased by 7-fold in obesity. Genes significantly down-regulated in obese subdermal fat included SLC27A2 (acetyl-CoA-synthase; 10-fold) and C6-complement (5-fold).

By QTPCR in subdermal fat from obese subjects, the increased expression of genes encoding proteins important in inflammation and immune function was confirmed, including genes encoding proteins that determine accumulation of immune cells in adipose tissues such as CD52, the high affinity immunoglobulin gamma FC receptor (FCGR1β), CCL3, CZXCL8 (interleukin 8) and CLEC7A, a pattern recognition receptor found in monocytes and other myeloid cells (Figure 7). IL17F, a member of the IL17 family, also was specifically increased in subdermal fat from the obese as compared to non-obese individuals.

Expression pathway analysis (Figure 8) showed upregulation of several inflammatory immune pathways including the T-helper 2, dendritic cell maturation and inflammatory signalling pathways further indicating profound effects of obesity on inflammation in subdermal fat. Dramatically lower in the obese subdermal fat was the LXR/RXR activation pathway.

**Figure 7.**
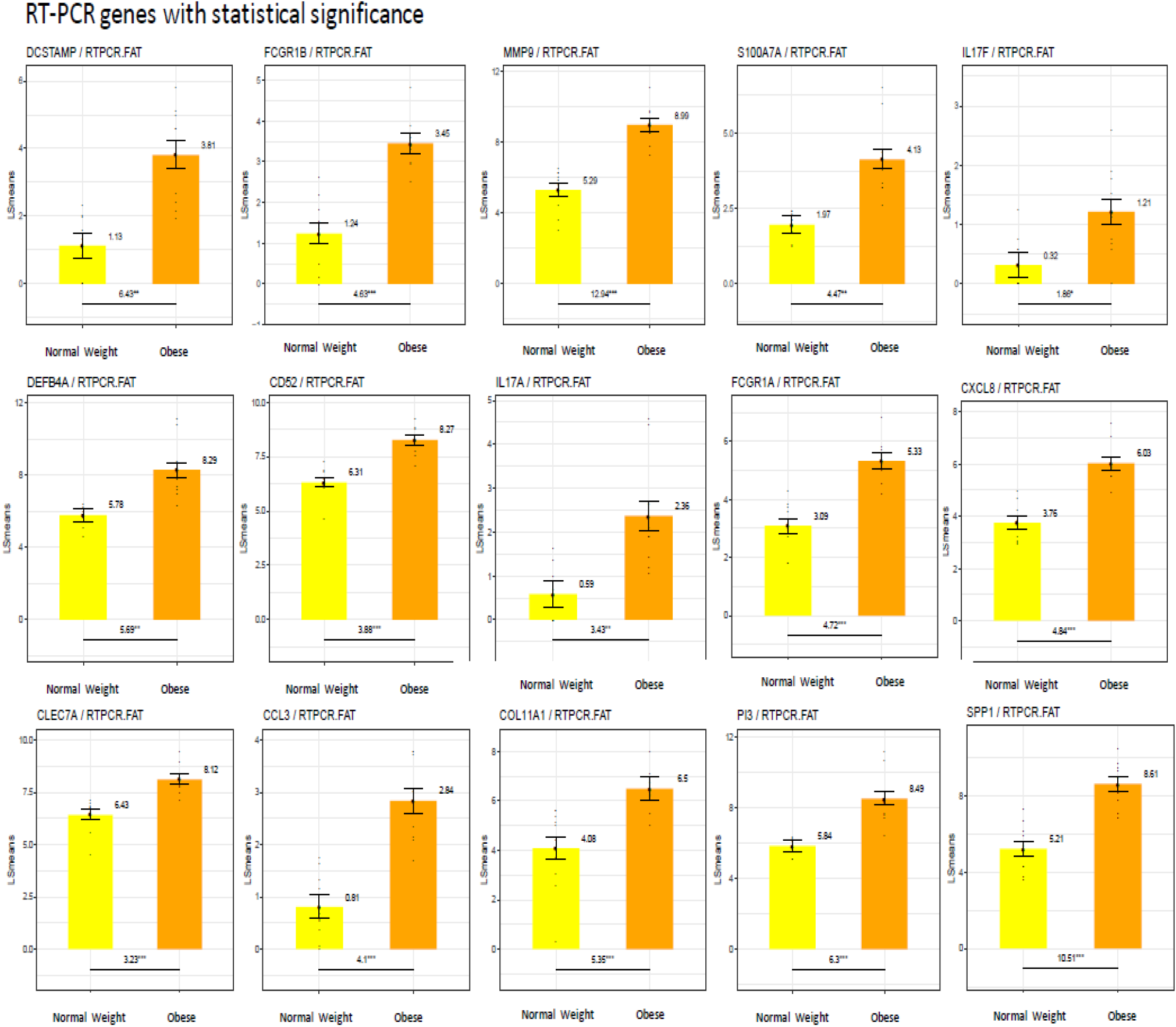
Differences in gene expression by RT-PCR between the subdermal fat of obese and non-obese subjects. LS means of gene expression by RT-PCR showing significant differences as *p<0.05; **p<0.01; ***p<0.001

**Figure 8.**
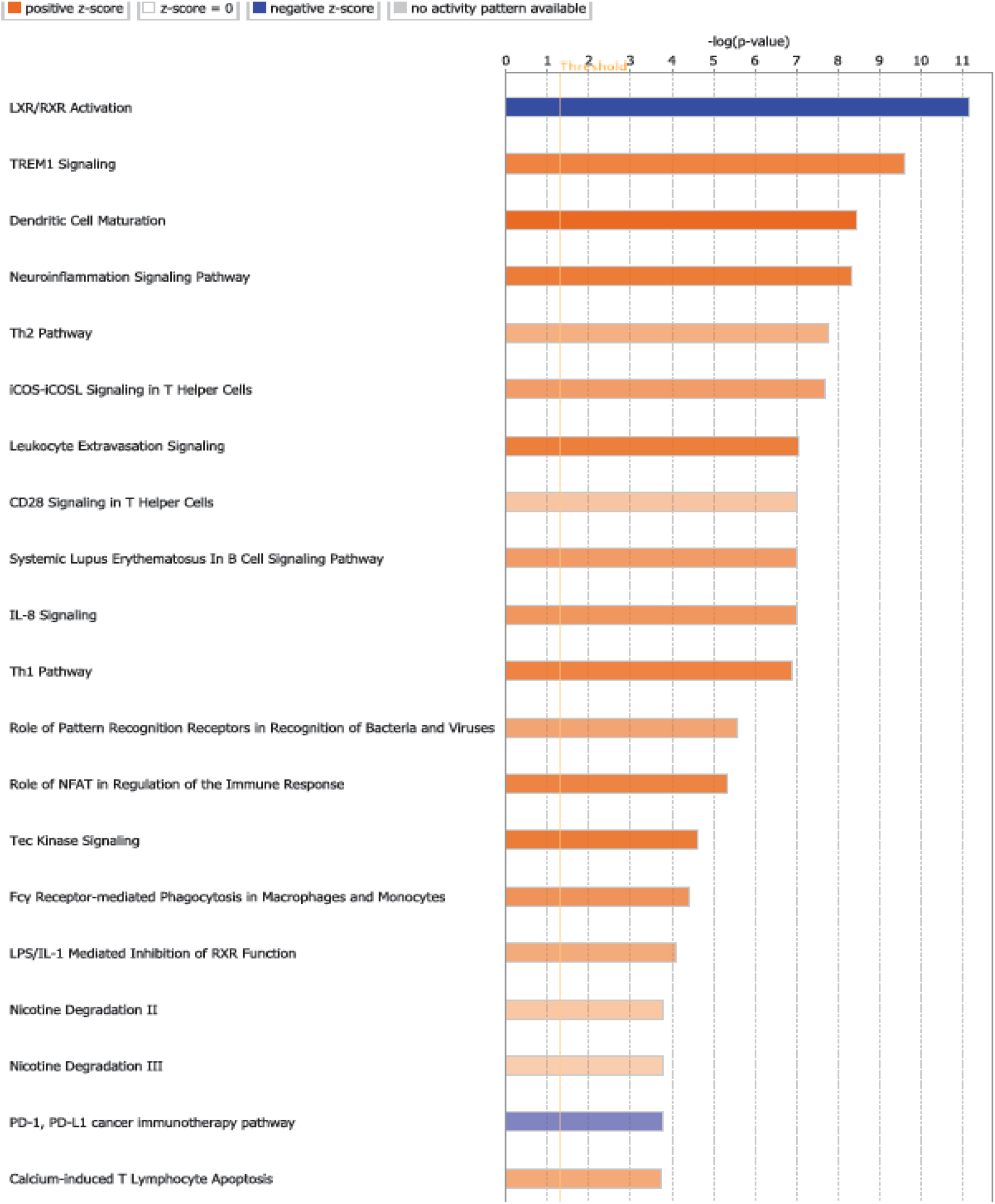
Gene expression pathway analysis between the subdermal fat of obese and non-obese subjects. Data shown from analysis of all differentially expressed genes. Log value shown in blue = negative *z* score Log value shown in orange = positive *z* score.

### Gene expression analysis by race

Two subgroups were observed in the gene expression profiles of the skin based on the subject’s race. Using self-reported data and skin colour as criteria, the data from African Americans was analysed separately from the data from Caucasians. This analysis showed striking differences in this very small group of subjects. A heatmap of the 50 most differentially expressed genes in skin from obese subjects divided by race is shown in Figure 7. No clear difference in gene expression is seen by race in the non-obese subjects. In contrast, gene expression clearly differed between obese African Americans and obese Caucasians (Figure 9A). The marked difference between obese African American and Caucasian subjects in gene expression is illustrated in the PCA plot in Figure 9B being responsible for the largest source of variation (PC1).

**Figure 9.**
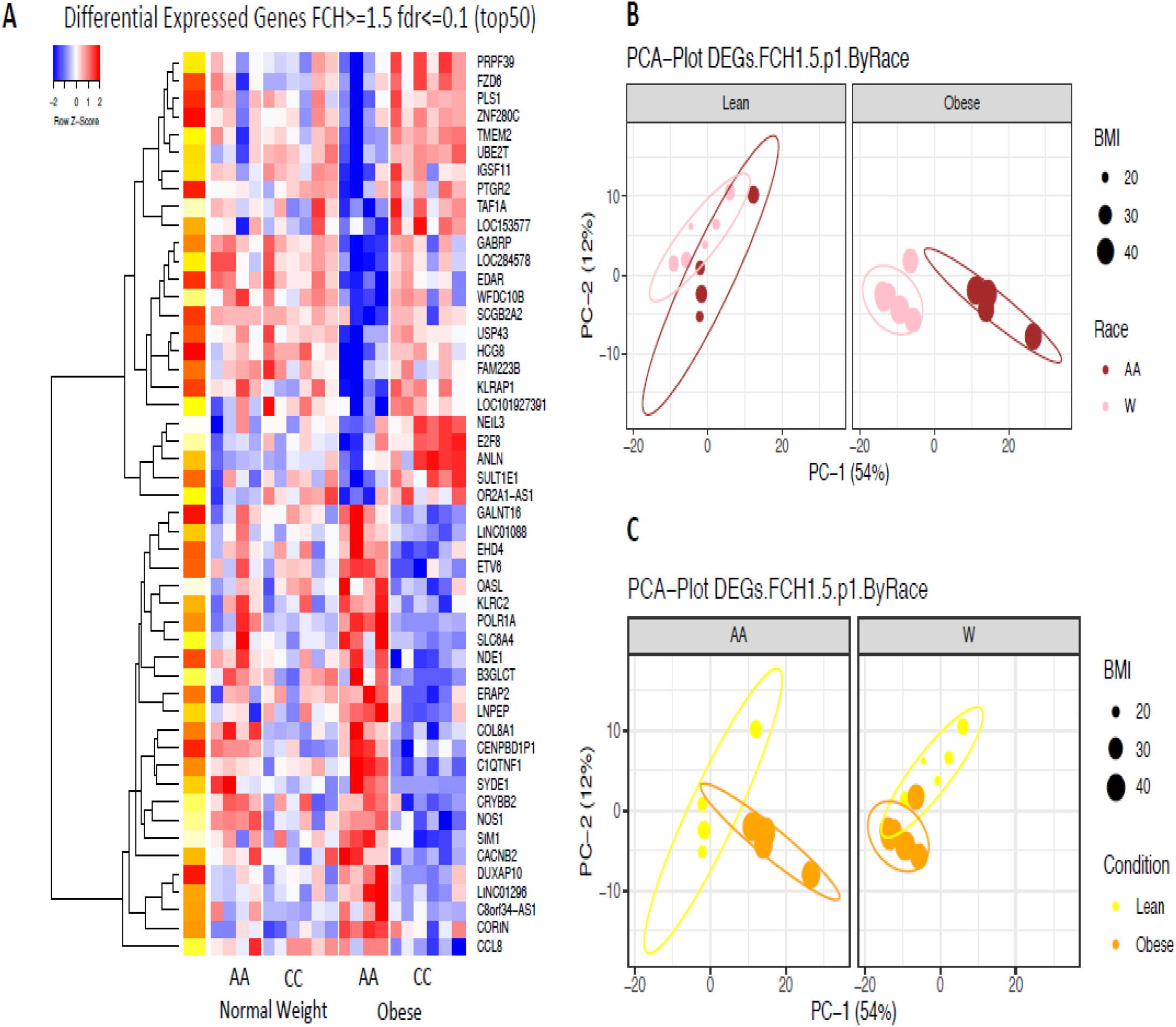
Differences in gene expression between the skin of African American and Caucasian subjects A. Heat map of the 50 most differentially expressed genes in skin of African American and Caucasian subjects with an FCH>=1.5; fdr<=0.1 B. PCA plot of differentially expressed genes in skin of obese and non-obese African American and Caucasian subjects with an FCH=1.5. Left side of plot indicates differences in gene expression by race in non-obese subjects. Right side of plot indicates differences in gene expression by race in obese subjects. C. PCA plot of differentially expressed genes in skin of obese and non-obese subjects by race with an FCH=1.5. Left side of plot indicates differences in gene expression between obese and non-obese African American subjects. Right side of plot indicates differences in gene expression between obese and non-obese Caucasian subjects.

A list of genes whose expression significantly differed between the obese African Americans and the obese Caucasians is shown in Tables S6 & S7. The expression of SLC6A4, a serotonin transporter, CORIN, and COL8AI – a collagen gene encoding a protein that is dysregulated in atopic eczema, was higher in African Americans, while the expression of SCCB2A2, the secretoglobin expressed in skin sweat glands, and CFTR was expressed higher in Caucasians.

Comparing the skin of obese African Americans to obese Caucasians by QTPCR, the former showed significantly lower expression of MYBCPI (5.16-fold) and PROM1 (4.3-fold) and CFTR (Figure 10). In contrast, there was a small increase in the expression of CORIN (2.2-fold), a gene encoding the atrial naturalistic peptide converting enzyme and BMP2 (1.33-fold) also present in the skin compared to obese Caucasians.

**Figure 10.**
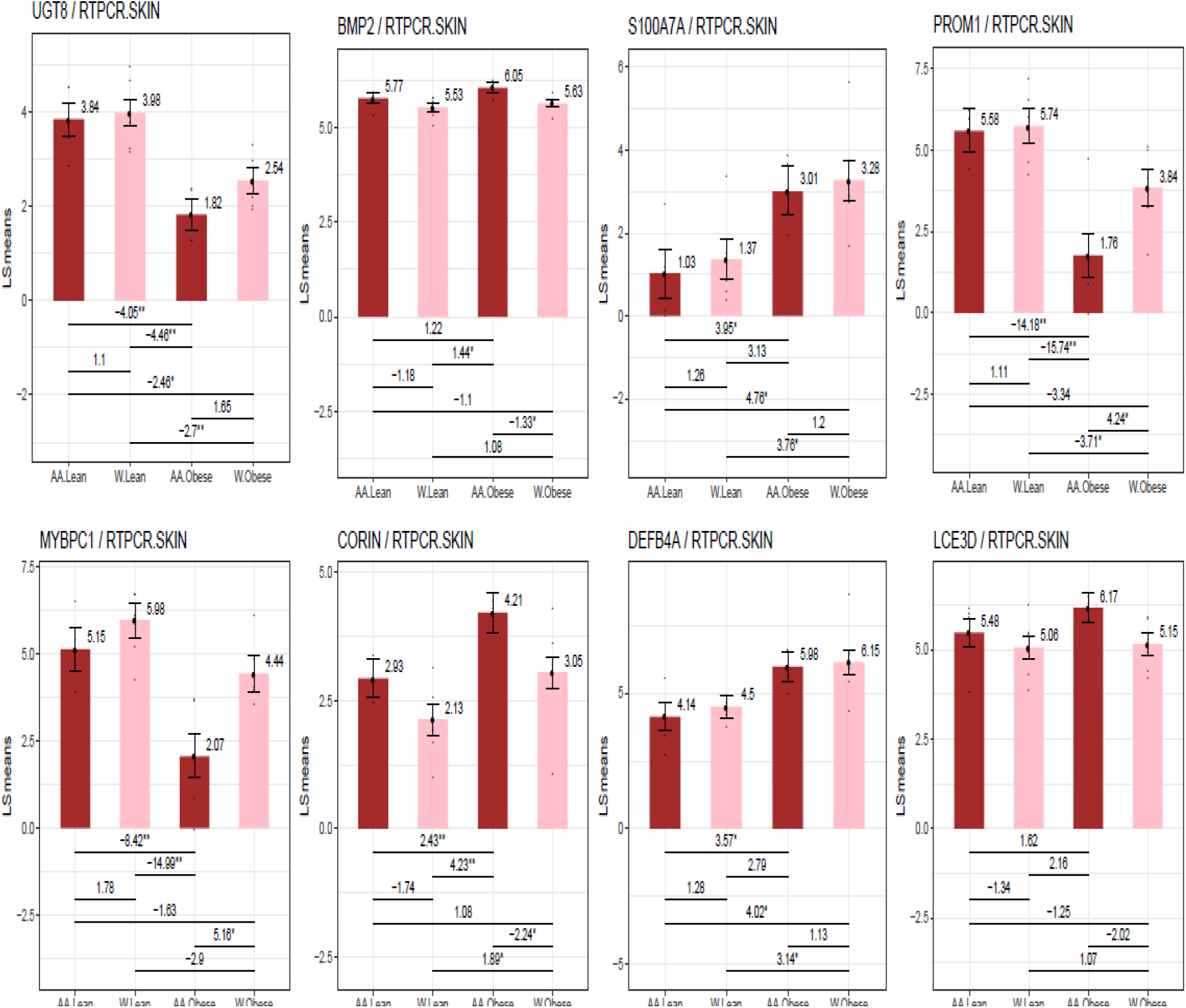
Differences in gene expression by RT-PCR between the skin of obese and non-obese African American and Caucasian subjects. LS means of gene expression by RT-PCR showing significant differences as *p<0.05; **p<0.01; ***p<0.001.

Pathway analysis (Figure 11) found no race-related differences in the non-obese samples. However, in the obese, there was markedly reduced expression of oestrogen mediated s-phase entry pathway, aryl hydrolase receptor signalling, pathway, and cell cycle regulation through cyclins pathways in the African Americans compared to the skin of the Caucasian group.

**Figure 11.**
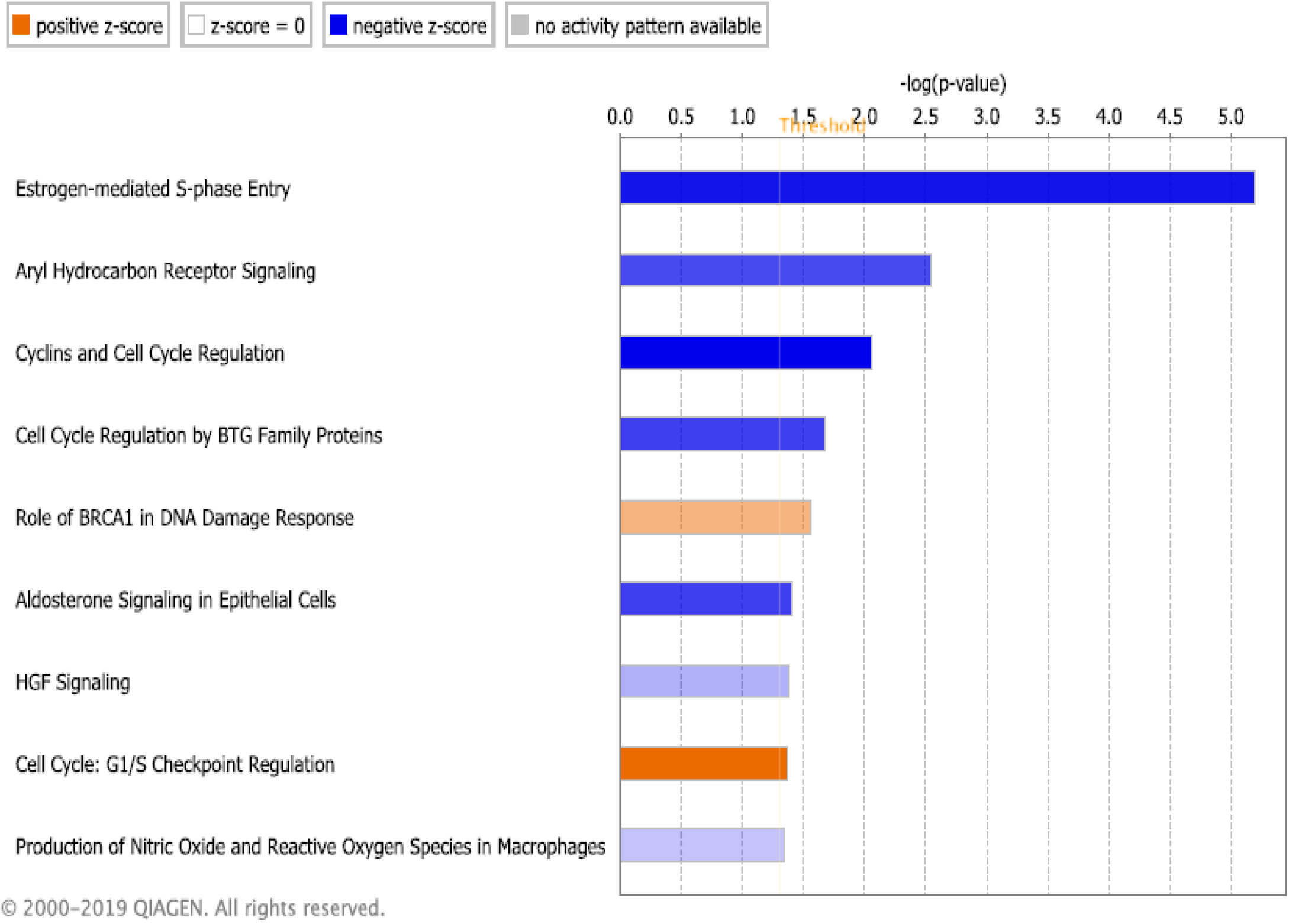
Gene expression pathway analysis between the skin of obese African American and obese Caucasian subjects. Data shown from analysis of all differentially expressed genes. Log value shown in blue = negative *z* score Log value shown in orange = positive *z* score.

**Figure 12.**
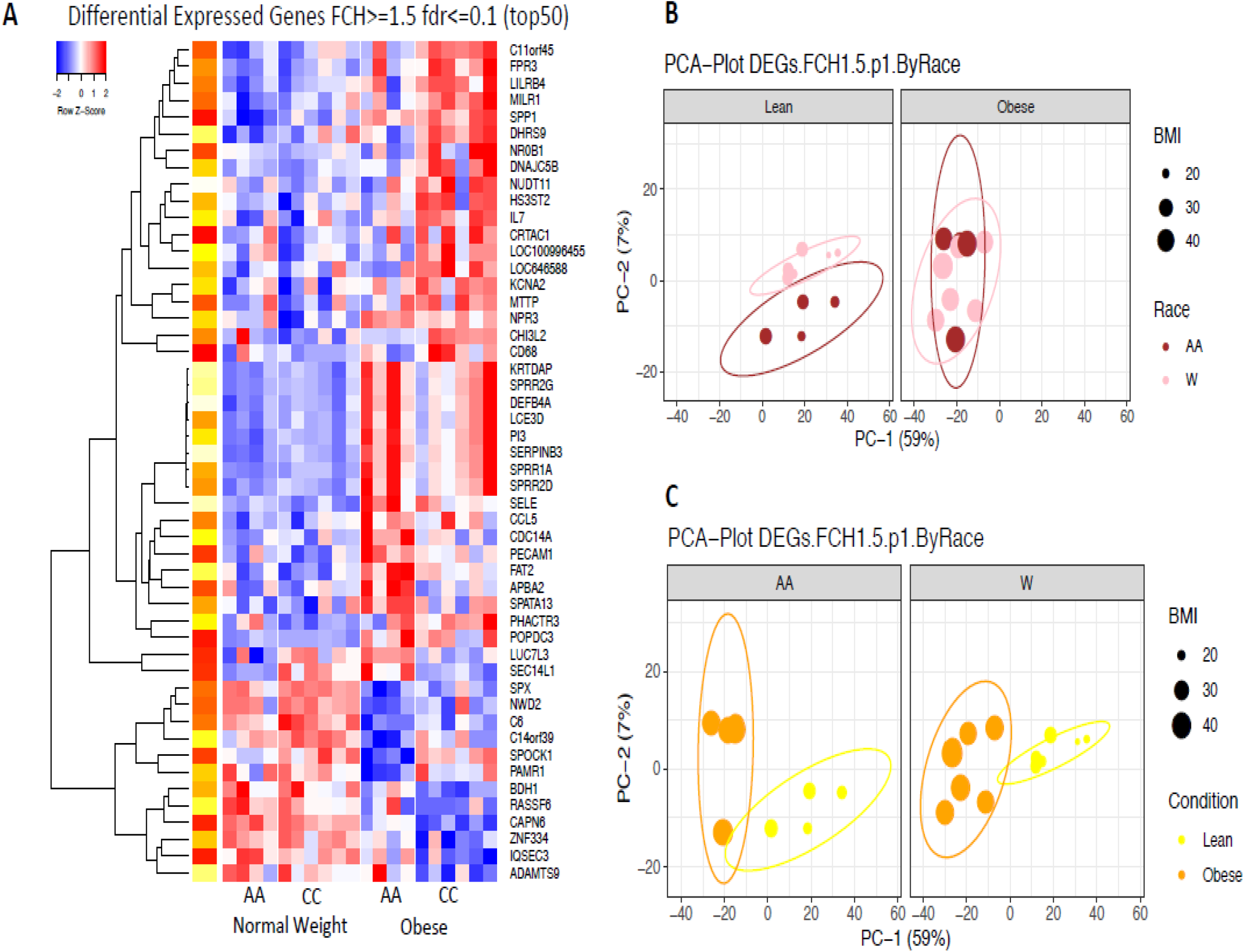
Differences in gene expression between the subdermal fat of African American and Caucasian subjects A. Heat map of the 5G most differentially expressed genes in subdermal fat of African American and Caucasian subjects with an FCH>=1.5; fdr<=0.1 B. PCA plot of differentially expressed genes in subdermal fat of obese and non-obese African American and Caucasian subjects with an FCH=1.5. Left side of plot indicates differences in gene expression by race in non-obese subjects. Right side of plot indicates differences in gene expression by race in obese subjects. C. PCA plot of differentially expressed genes in subdermal fat of obese and non-obese subjects by race with an FCH=1.5. Left side of plot indicates differences in gene expression in obese and non-obese African American subjects. Right side of plot indicates differences in gene expression in obese and non-obese Caucasian subjects.

**Figure 13.**
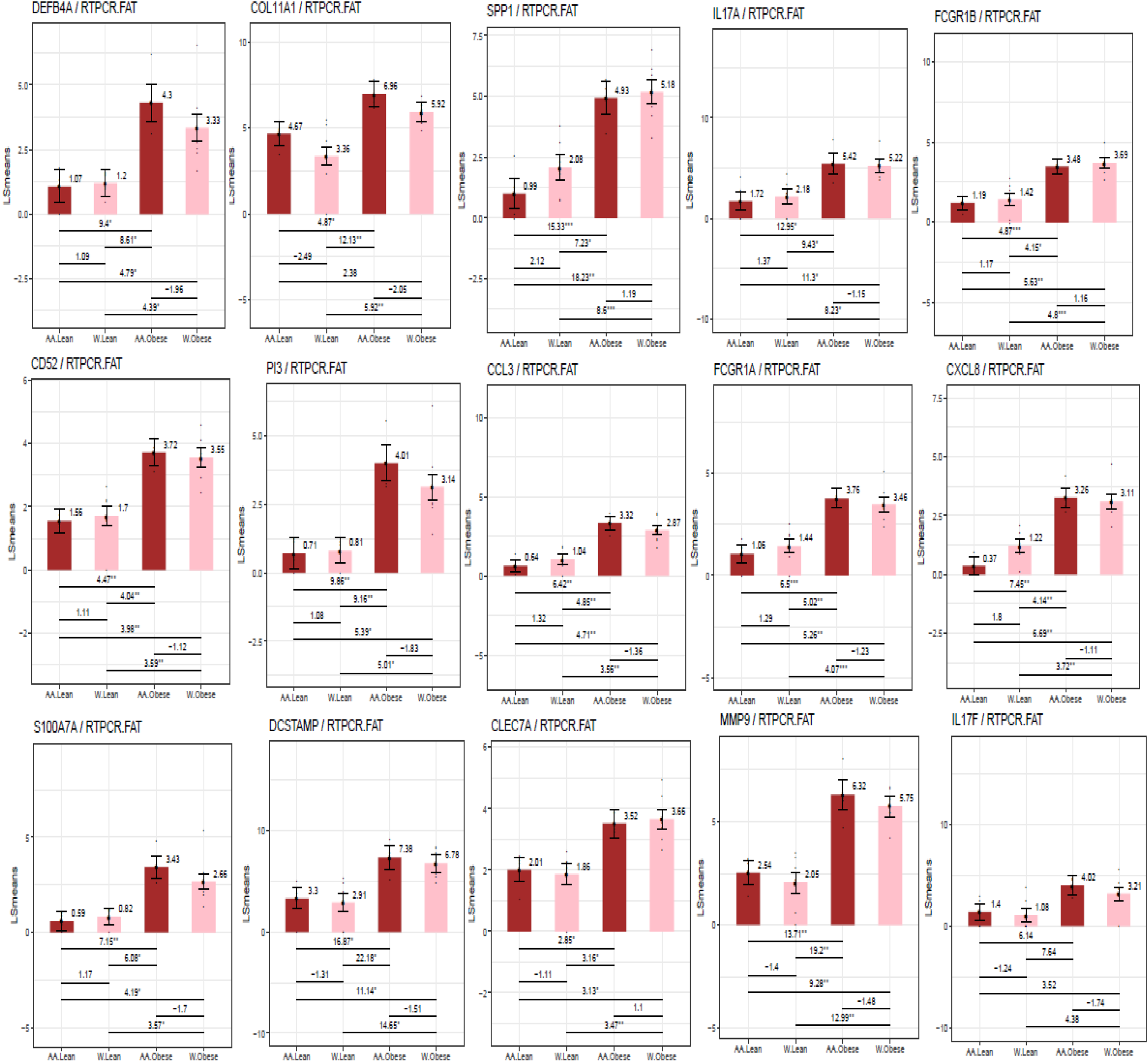
Differences in gene expression by RT-PCR between the subdermal fat of obese and non-obese African American and obese and non-obese Caucasian subjects. LS means of gene expression by RT-PCR showing significant differences as *p<0.05; **p<0.01; ***p<0.001.

**Figure 14.**
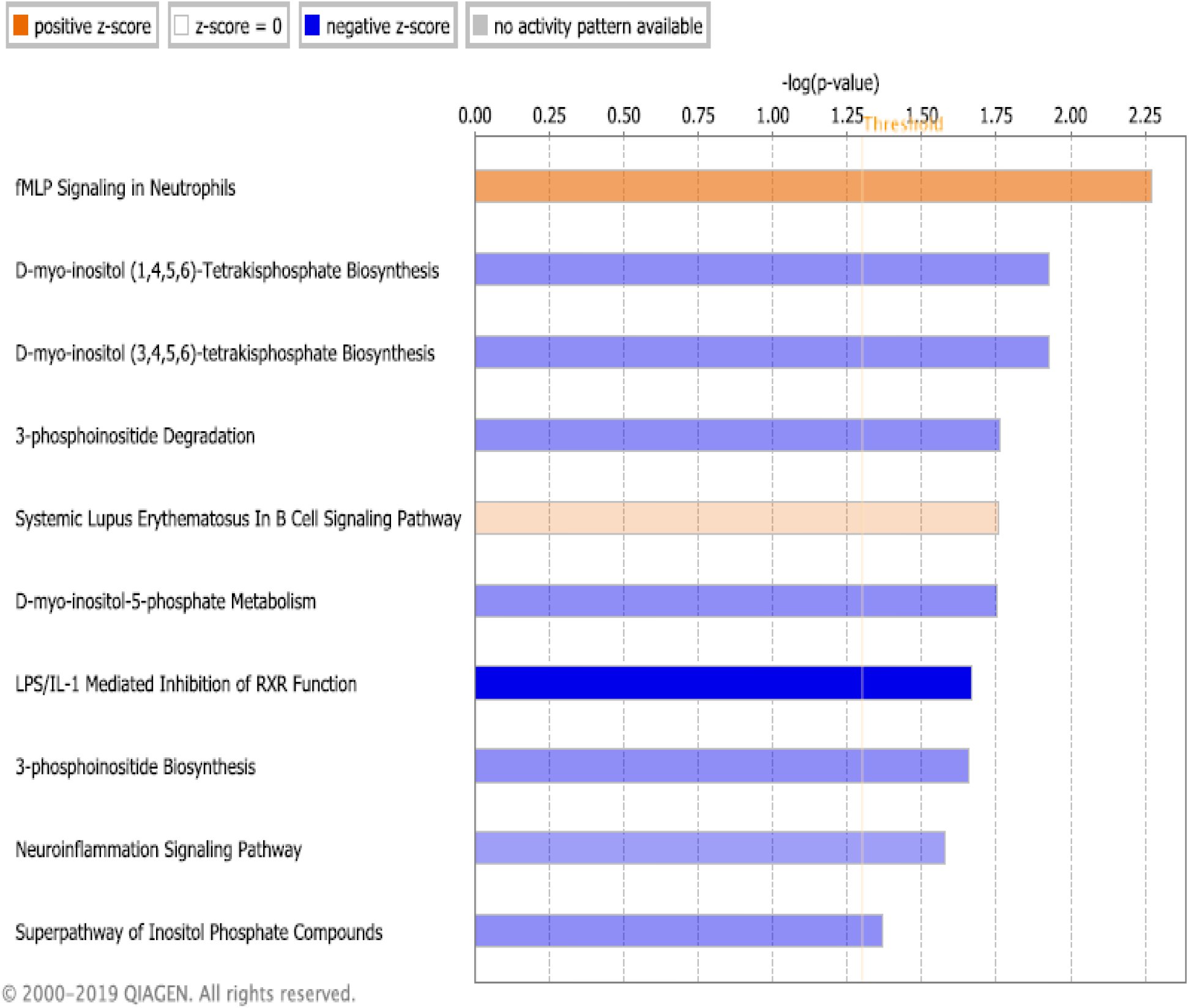
Gene pathway analysis between the subdermal fat of obese African American and obese Caucasian subjects. Data shown from analysis of all differentially expressed genes. Log value shown in blue = negative *z* score Log value shown in orange = positive *z* score.

### Microbiota analysis

We next examined whether the microbiota collected from skin swabs around the biopsy site differed between the obese and non-obese subjects. We generated taxonomic profiles for each sample using KrakenUniq and a database built from all available microbial species in RefSeq. We measured the total number of AMR genes detected in each sample by aligning reads to MegaRES.

Overall differences between groups were minor. No significant differences were noted in average taxonomic alpha diversity as measured by either Shannon’s entropy or richness between the groups. A PCA plot of the taxonomic profiles showed slight separation between obese and lean samples and slightly higher beta diversity for obese samples however these differences were minor (Figure 15A).

**Figure 15.**
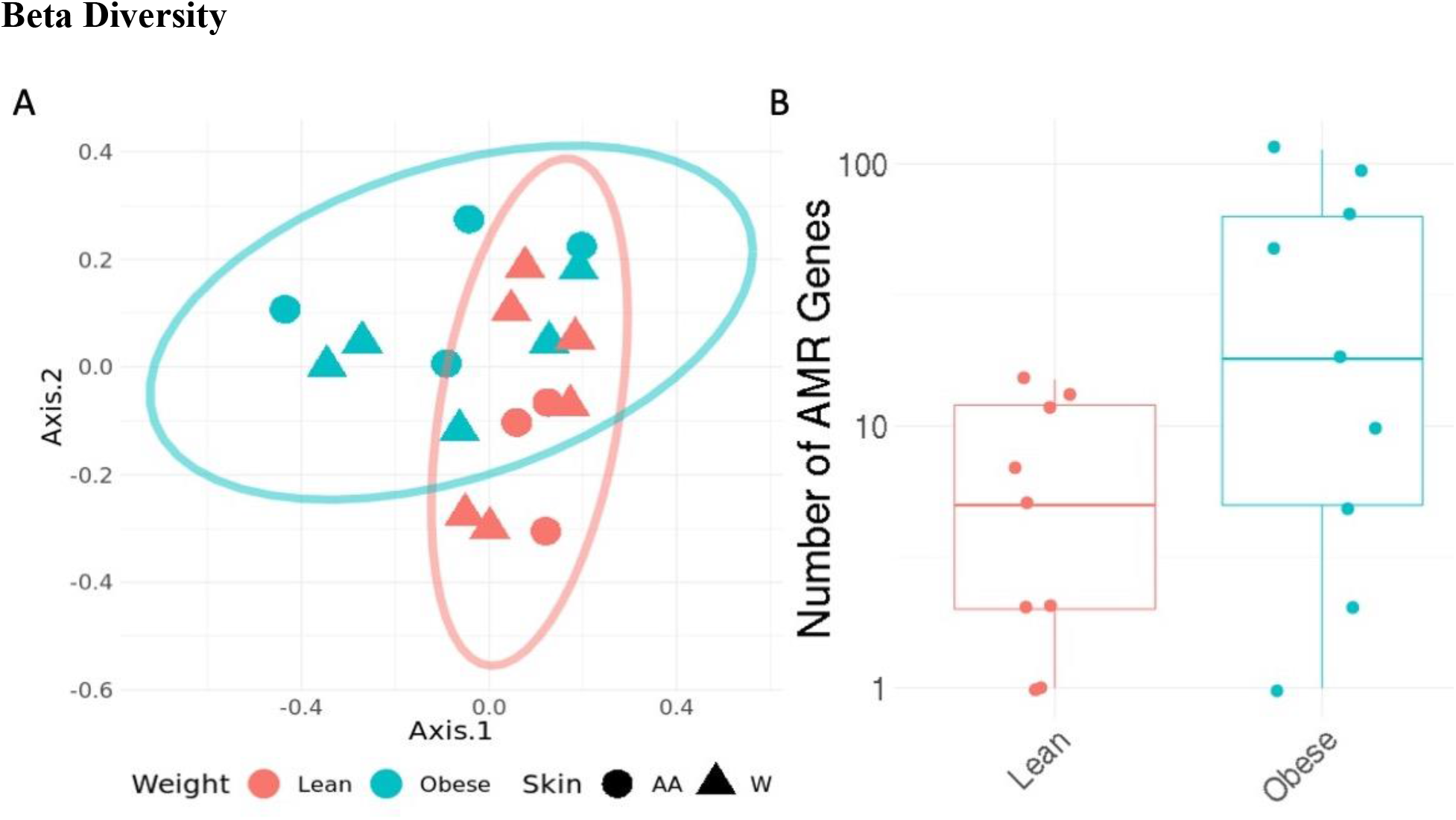
Metagenomic analysis of skin microbiomes A. PCA plot of taxonomic profiles for skin microbiomes from obese and non-obese subjects B. Total number of Antimicrobial Resistance Genes detected in samples (*p*=0.1; Wilcox test).

## Differentially Abundant Taxa

At the given sample size (*n* = 10) no taxa were identified as significantly differentially abundant after Benjamini-Hochberg correction. Before correction five taxa were significantly differentially abundant at *p* = 0.005 (Wilcox test). These five taxa were *Corynebacterium aurimucosum, Corynebacterium jeikeium, Corynebacterium urealyticum, Streptococcus salivarius*, and *Streptococcus sp. A12*. All five taxa were more abundant in samples from obese subjects on average.

### Metabolome analysis

As highlighted by the PCA analysis, no variation in the metabolites analysed by liquid chromatography-mass spectrometry from the skin swabs was observed between the obese and non-obese individuals (PCA supplemental Figure S1). Similarly, no race-related metabolic variation was observed. These results were further confirmed by the poor predictive ability of the orthogonal projection to latent structures-discriminant analysis (OPLS-DA) models comparing the two different racial groups.

## DISCUSSION

Few comprehensive reviews of skin changes occurring with obesity have been conducted despite over 40% of the US population being obese [20]. A broad review of the physiologic and clinical consequences and associations was published by Hirt et al in 2019 [21]. The authors include a discussion of circulatory and lymphatic changes which may enhance the frequency and severity of skin ulceration and provide a comprehensive review of skin disorders that can be associated with obesity, expanding on previous reviews [22] and studies in rodents [23].

In our study, the thickness of the epidermis did not differ between the obese and non-obese subjects. Our data contrasts with a Japanese study in which skin taken from women undergoing breast reconstruction showed a shift to greater epidermal thickness in the obese when compared to non-obese subjects, but the variation was very large [24]. In another study in obese subjects in which dermal structure in abdominal skin was visualized as echogenicity using ultrasound scanners, weight reduction decreased dermal thickness in about half of the subjects [25]. A previous study also observed no increase in skin thickness in obese subjects weighing up to 200 kg. [26]. In rats with obesity and diabetes mellitus, epidermal thickness was reported as lower than in normal weight animals [27]. Six months of caloric restriction in mice increased epidermal thickness almost twofold [28].

An important cutaneous manifestation of obesity is chronic skin ulceration which can result from pressure from the weight of the body on the perineum or back and from venous congestion and a reduced blood supply to the lower extremity [22]. Our data suggest potential molecular factors that could contribute to defects in wound healing including altered blood flow, and less mediators of the normal healing process.

Normal delivery of oxygen and nutrients are crucial for wound healing and a reduced blood flow could exacerbate skin ulceration. Impaired vasodilatation and vasoconstriction occur early during the development of obesity [29]. Although total blood flow is increased in obesity [30] and skin temperature can be higher [31] the microvasculature may be inadequate to maintain the viability of healing skin tissues. The skin microcirculation is maintained by appropriate responses to need in part by endothelin-1 activity which can alter blood flow [32]. Additionally, reduced skin blood flow in obesity could be exacerbated by inflammatory changes in adjacent adipose tissue [29]. Our studies showed reduced endothelin-1 gene expression in the skin of obese subjects and pronounced inflammation in the subdermal fat. Proinflammatory cytokines from such inflamed fat also stimulates S100A7 and DEfB4A expression and can cause endothelial dysfunction in local blood vessels in part due to inadequate endothelin-1 action [33].

The S100A7 gene showed the greatest differential upregulation in the skin of obese compared to non-obese subjects. This gene encodes a protein which functions in cell cycle proliferation and differentiation and strengthens the skin-tight-junction barrier. The pro-inflammatory cytokines IL 17 and IL22, which are secreted from obese adipose tissue, stimulate S100A7 expression in keratinocytes. [34].

In contrast, the beta-adrenergic receptor pathway was the most downregulated in the skin of obese subjects in our study. Keratinocytes express only beta-2 adrenergic receptors for the catecholamines epinephrine and norepinephrine [35]. Skin wounding generates epinephrine which binds to its receptors resulting in excessive adrenergic activity and reduced skin wound healing by altering keratinocyte migration [36]. Defects in beta adrenergic activation are associated with the severity of atopic dermatitis and psoriasis [37]. Whether these changes affect keratinocyte functions enough to contribute to defective healing of skin wounds in obese skin has not been established.

Increased inflammatory-immune activity has been extensively described in obese subcutaneous and visceral fat [38]. However, several observations suggest that subdermal fat differs in molecular composition and in function from adipose tissue in other sites [39].

Increased inflammation in dermal fat also may result in altered skin microvascular reactivity as shown in mice [23] and in skin temperature in humans [31]. Clinically, obesity may precede psoriasis at least in children [40]. The skin barrier is maintained by normal keratinocyte proliferation but the expression of several genes encoding proteins determining optimal proliferation were altered in the skin of obese subjects. As an example, the gonadotrophin releasing hormone (GNRH) pathway was significantly reduced in the skin of the obese compared to non-obese subjects [41]. Although GNRH is a central regulator of reproductive functions, it also may help regulate cell proliferation and cell motility [42].

Subdermal fat (dWAT) of obese subjects showed a generalized increase in inflammatory-immune activity compared to the non-obese including changes in genes encoding proteins that induce leucocyte attraction, increased activity of the TH1 pathway and in dendritic cell activation. Furthermore, there was evidence of a major increase in signalling by TREM a receptor for MCP and TNF [43, 44] and in osteopontin encoding a protein that increases interferon gamma and IL-12 activity.

There is extensive literature showing that obesity is accompanied by intense inflammatory-immune changes in subcutaneous and visceral fat [38] but subdermal fat has received little attention. Adipocytes stimulate follicle regeneration [45], are recruited to skin wounds [46], have been described as a 1 mm thick layer [47] or as pilosebaceous units (dWAT) with a high turnover rate, an important factor in skin scarring [48]. Adipocytes secrete antimicrobial peptides and cytokines that signal hair follicles [44] to activate and modulate rates of hair growth and facilitate wound healing from injury and infection [50]. These data demonstrate the unique plasticity of these adipocytes in maintenance of skin homeostasis [51].

There were significant differences between the skin of obese African Americans compared to obese Caucasians. Remarkably, this was evident even though only 4 paired obese and 4 non-obese African Americans and 6 paired obese and non-obese Caucasian subjects were studied. Differences occurred in epidermal thickness, oestrogen metabolism, aquaporin and hydrocarbon receptor signalling and in gene expression encoding cadherins, platelet activating factors and solute carriers, including CFTR.

We found a trend (p = 0.08) for a wider epidermis in the African American subjects than in Caucasians. Some [51, 52] but not all [53] previous studies have also shown that African Americans have increased skin thickness as well as a wider cornified layer, greater desquamation [54] and differences in skin lipids but a lower water content [55]. African Americans skin is said to age at a slower rate with less wrinkling and sagging, better barrier function but greater sensitivities to exogenous chemicals [56] and poorer wound healing rate [57].

In our study, skin gene expression in non-obese African Americans and Caucasians showed very few differences but the obese groups differed widely. In African Americans, the expression of the SLCA4 gene encoding a serotonin transporter (SERT), which can enhance skin sensitivity, was increased as was 5-hydroxytryptophane which stimulates keratinocyte

Oestrogens have many beneficial effects on skin physiology since the skin of post-menopausal women suffers defects which leads to “aging skin” changes and oestrogens prevent or reverse many of these effects [58]. Thus, the increase in oestrogen pathway gene expression may be important to explain why the skin of African American ages more slowly than that of Caucasians. Oestrogens can stimulate greater fibroblast proliferation with thickening of the skin [59] and can act as an antioxidant [60].

The expression of the aryl hydrocarbon receptor (AHR) signalling pathway was lower in African Americans than Caucasians. The action of AHR on keratinocytes is part of the ultraviolet light stress response [61]. In the absence of the AHR, UVB exposure induces less production of melanocytes. Possibly, skin tanning and pigmentation may involve AHR signalling [62] and that black skin requires less AHR activity. However, reduced AHR activity may, at the same time, enhance inflammatory skin conditions such as psoriasis [63].

Our study in African Americans showed an about 4-fold lower expression of the CFTR gene that encodes the cystic fibrosis transmembrane conductor regulator, a channel protein that regulates chloride transport across membranes. Abnormalities of this gene, which has over 2000 mutations, is the cause of human cystic fibrosis. The incidence of cystic fibrosis is lower in African Americans (1 in 15,000) compared to about 1 in 2500 in Caucasians. One reason for a lower incidence may be that mutation testing that is routinely performed at birth in developing countries is based on the mutations seen in patients in the United States [64] and that the mutation spectrum in African nations differs significantly from that seen in Caucasians in the United States and in Europe [65]. The CFTR protein also functions in normal skin since defects in skin wound healing occur when CFTR protein function is genetically deranged [66]. CFTR expression is found in multiple layers of the skin [67] even though the skin is not an epithelial transport system. *In vivo* and *in vitro* studies have shown CFTR upregulation in early and late wound healing in CFTR −1-knock out mice [68]. Whether reducing CFTR function contributes to delayed wound healing in obesity remains to be determined.

Regulation of body temperature is by sweating; water is excreted via sweat glands, evaporates, and sodium is reabsorbed by epithelial sodium channels (ENaC). The main physiologic regulator for this process is the renin aldosterone system whereby aldosterone binds to mineralocorticoid receptors which are expressed in keratinocytes. In cystic fibrosis, absorption of salt is impeded despite a functional ENaC, so that sweat sodium is poorly reabsorbed. ENaC and CFTR appear to localize in “channels” in sweat glands.

In obesity and African Americans there is evidence of differences in skin moisture with increase in trans epidermal water loss(70) and capacitance resulting in dryer, rougher and more scaly skin [22]. Factors in our study that may be responsible include lower expression of aquaporin which is 5.8-fold greater in Caucasians and changes in ion transport. Aquaporins represent membrane proteins that function to form water channels and are involved in water homeostasis and skin hydration [69]. Although the impact of this relative increase in aquaporin-5 gene expression in Caucasians may have on the biology of the skin is presently unclear, skin hydration and wound healing [70] do differ between the skin of Caucasian subjects and African Americans.

The most detailed previous evaluation of differences between the skin biology in African Americans and Caucasians were performed with *in vitro* reconstructed skin [54] using isolated primary cells, keratinocytes and dermal fibroblasts from the same donor in long-term culture [71, 72]. The authors examined skin types II, II, IV from four African American and Caucasian donors to evaluate structure using immunohistochemistry, microarrays for gene expression and proteomics. They described increased convolutions in African American skin like that seen in our studies *in vivo*, but also no difference in skin thickness. In addition, they reported about 425 genes that were differentially expressed that function predominantly in lipid and filaggrin processing. Similar to our findings, they described significant differences in the expression of SLC44A5, but also genes and proteins involved in terminal differentiation and concluded that the dermal-epidermal junction length was 3-fold greater in African Americans than in Caucasians providing an expanded exchange area between these layers of the skin.

We examined skin microbiota derived from swabbing the surface of the skin adjacent to the site of the skin biopsy but found only minor differences between obese and non-obese subjects in the structure of these microbiota. A PCA plot suggested that microbiota from non-obese subjects were more tightly clustered than from the obese but there was much overlap. Skin surface microbiota interact closely with adjacent skin [73], are fundamental to skin physiology and immunity and are important as secondary factors that can determine the course and treatment of primary skin disorders [74]. Skin infections such as folliculitis and erysipelas are more common in obesity. We have found only one study in the literature that attempted to determine effects of obesity on the skin microbiome of “healthy” individuals. That study used data derived from the American Gut Project which gathered oral, faecal, and skin samples from the public at large [75]. Skin samples from 81 self-reported obese, 80 underweight and 580 normal weight individuals from around the world suggested that obesity was accompanied by an increase in Corynebacterium species [76]. Vongsa [77] and Rood [78] reported that obesity led to no significant changes in the distribution of skin microbiota in premenopausal or pregnant women. Since we only examined a small number of subjects it was not possible to draw conclusions on effects of obesity upon skin microbiota. However, the data we have made it unlikely that microbiota played a crucial role in the molecular changes within the skin that we observed between the obese and non-obese or between African Americans and Caucasian subjects.

No significant differences were found in the metabolic profiles of the skin swabs collected close to the site of the biopsy in any of our groups. Julia Oh [79] compared metabolites derived from differing skin sites and emphasized the marked variation between individuals. Others explored the potential importance of measuring sweat metabolites in studies of the skin [80]. None of these studies focused specifically on obesity or the ethnicity of the subjects.

In summary, we acknowledge that the number of subjects in our study was very small, but the data was reproducible by differing techniques. We show that the non-exposed abdominal skin of obese post-menopausal female subjects differs from that of age and race matched non-obese post-menopausal women. Although epidermal thickness of the 2 groups did not differ, there was marked upregulation of the S100A7 gene that encodes proteins that can alter skin oxygenation and can contribute to poor wound healing of skin ulcers that can afflict obese skin. Subdermal fat showed marked upregulation of genes encoding inflammatory and immune altering proteins like what has been described in deep subcutaneous and visceral adipose tissues in obesity.

We show evidence that points to differences between the skin of African American and Caucasian subjects. The molecular changes observed suggested impairment of the water and solute transport as well as aryl hydrocarbon receptor action in the skin of African Americans. Neither skin surface microbiota nor metabolites were altered to suggest their involvement in the skin changes that we observed, but the subdermal adipose tissue inflammatory-immune activity is likely to have been important.

## Data Availability

All data is provided in the manuscript submitted. The gene expression data has been downloaded into the GEO repository. The accession number is pending.

https://www.ncbi.nlm.nih.gov/geo

## Acknowledgements

We thank the participants and clinical research staff who made this clinical study possible.

We thank Pat Gilleaudeau, FNP for assistance with the punch biopsy procedure

We thank Inna Cueto for staining and photographing the specimen slides

We thank Chandrima Bhattacharya for assistance with microbiome analysis

This research was supported in part by RUCCTS grant # UL1TR001866 from the National

Center for Advancing Translational Sciences (NCATS), National Institutes of Health (NIH)

Clinical and Translational Science Award (CTSA) Program (JMW).

JOA was funded by the Center for Diseases of the Digestive System and Sackler Center for Bionutrition at Rockefeller University, AHA 17-SFRN33520045 and K08-DK117064 (NIH) at NYU Langone Health.

## Author Contributions

JMW designed and conducted the study

SG evaluated the data and did the statistical analysis

JOA helped to evaluate and write the manuscript

CM helped to analyse and write up the microbiota data

DD helped to analyse and write up the microbiota data

SZ helped run and analyse the metabolite data

JS helped design, analyse, and write up the metabolite data

JK helped to analyse and write up the data

JLB helped to analyse, interpret and write up the data

PRH designed, analysed, interpreted, and wrote up the data

## Author approval

All authors have read the manuscript and approved its contents.

## Disclosures

All authors have completed the ICMJE uniform disclosure form at www.icmje.org/coi_disclosure.pdf and declare: no support from any organisation for the submitted work.

JMW received research grants from RUCCTS grant # UL1TR001866 from the National Center for Advancing Translational Sciences (NCATS), National Institutes of Health (NIH) Clinical and Translational Science Award (CTSA) Program.

JOA received research grants from the Center for Diseases of the Digestive System and Sackler Center for Bionutrition at Rockefeller University, AHA 17-SFRN33520045 and K08-DK117064 (NIH) at NYU Langone Health

## Data Availability Statement

All data referred to in the manuscript is freely available and can be found in the body of the manuscript or in the supplemental material. Gene expression data was deposited into Gene Expression Omnibus (GEO). The GEO number is pending.

## Supplemental Materials

### Methods

#### Skin microbiome

The DNA extraction protocol was adapted from the Maxwell RSC Buccal Swab DNA kit (Catalogue number AS1640: Promega Corporation, Madison WI). Briefly, 300 μl of lysis buffer and 30 μl of Proteinase K was mixed and added to each swab tube. Swab tubes were then incubated for 20 minutes at 56 C using a Thermo Fisher water bath, removed from the tubes, and fluid was transferred to well #1 of the Maxwell RSC Cartridge. The swab head was centrifuged using a ClickFit Microtube (Cat. # V4741), and extracted fluid was added to the corresponding well of Maxwell Cartridge, and eluted in 50 μl of provided elution buffer.

Extracted DNA was taken forward to the Nextera Flex protocol by Illumina. Briefly, 30 μl of extracted DNA was taken into library prep protocol and run with 12 cycles of PCR. Libraries were cleaned up with a left sided size selection, using a bead ratio of 0.8x. The right sided size selection was omitted. Libraries were then quantified using a Thermo Fisher Qubit Fluorometer and an Advanced Analytical Fragment Analyzer. Libraries were sequenced on an Illumina HiSeqPE 50×2 at the Weill Cornell Epigenomics Core.

All bioinformatic analysis was performed on Weill Cornell Medicine’s Athena compute cluster, a high-performance grid compute system. Secondary analysis was performed on a Linux and MacOS systems. Unless otherwise noted programs were run with default settings.

Raw sequence data were processed with AdapterRemoval (v2.17) to remove low quality reads and reads with ambiguous bases [1]. Subsequently reads were aligned to the human genome (hg38, including alternate contigs) using Bowtie2 (v2.3.0, fast preset) [2]. Read pairs where one or both ends mapped to the human genome were separated from read pairs where neither mate mapped. Read pairs where only one mate mapped were discarded. Hereafter, we refer to the read sets as human reads and non-human reads.

Taxonomic profiles were generated by processing non-human reads with KrakenUniq (v0.3.2) with a database based on all draft and reference genomes in RefSeq Microbial (bacteria, fungi, virus, and archaea) ca. March 2017. KrakenUniq identifies k-mers that are unique to taxa in a database. Reads are broken into k-mers and searched against this database. Finally, the taxonomic makeup of each sample was given by taking the proportion reads which were assigned to each clade. KrakenUniq counts the number of unique marker k-mers assigned to each taxon and we filtered taxa with fewer than 512 unique markers [3].

We performed differential abundance testing over microbial species using the ALDEx2 R package. ALDEx2 performs variance stabilization read counts using a centred log ratio transformation that models samples as Dirichlet-Multinomial distributions over taxa then compares taxonomic abundances across groups [4]. Comparison of abundances across groups was done with a Wilcoxon rank sum test and Benjamini Hochberg Correction for multiple hypothesis testing.

Dimensionality reduction of taxonomic profiles was performed with Principal Coordinates Analysis based on a matrix of Jensen-Shannon Divergences (JSD) between samples. Analysis of inter-sample (beta) diversity was performed using the same matrix of JSD. Intra-sample (alpha) diversity was measured by finding Shannon’s Entropy of the taxonomic profile and by counting the total number of species identified in each sample (richness). Shannon’s entropy accounts for the relative size of each group in diversity estimation and is defined as 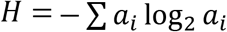 where *a_i_* is the relative abundance of taxa *i* in the sample.

We generated profiles of antimicrobial resistance genes using MegaRes (v1.0.1) [5]. To generate profiles from MegaRes, we mapped non-human reads to the database using Bowtie2 (v2.3.0, very sensitive presets). Subsequently, alignments were analysed using Resistome Analyzer (commit 15a52dd) and normalized by total reads per sample and gene length to give RPKMs. MegaRes includes an ontology grouping resistance genes into gene classes, AMR mechanisms, and gene groups.

#### Skin metabolome

The skin was swabbed using two sterile, saline-moistened culture swabs and immediately frozen at −80°C. Swab heads were removed and placed in 1 mL methanol: water (1:1). Following sonication (30 mins), 1 mL of isopropanol was added, and the solution was spun at 10,000 *g* for 30 mins. The swab was removed, and the samples were dried using a vacuum concentrator operating at 30 °C. Prior to UPLC-MS analysis, samples were reconstituted in 50 μL of HPLC-grade water, sonicated for 20 minutes and transferred to vials for analysis.

A Waters 2777C sample manager (Waters Corp., Milford, MA, USA) was used for sample handling. This was equipped with a 25 μL Hamilton syringe, a 2 μL loop used for full-loop injections of prepared sample, and a 3-drawer sample chamber maintained at 4 °C with a constant flow of dry nitrogen gas to prevent the build-up of condensation. The LC component was an ACQUITY UPLC (Waters Corp., Milford, MA, USA) composed of a binary solvent manager and column heater/cooler module. Metabolic profiles were acquired using reversed-phase chromatography. Water and acetonitrile, each supplemented with 0.1 % formic acid (mobile phases A and B, respectively), were selected for the mobile phase. A 2.1 × 150 mm HSS T3 column was used at 45 °C with a mobile phase flow rate of 0.6 mL/min. This generated a maximum pressure of ~12,000 psi in a water/acetonitrile gradient. After a 0.1 min isocratic separation at initial conditions (99% A), a linear gradient elution (99% A to 45% A in 9.9 min) proceeded, followed by a quicker gradient (45% A to 0% A in 0.7 min) to final conditions. The mobile phase flow rate was simultaneously increased to 1.0 mL/min in the latter stage to facilitate faster column washing. The MS component comprised a Xevo G2-S QToF MS (Waters Corp., Manchester, UK) coupled to the UPLC via a Zspray electrospray ionization (ESI) source. The cone gas flow was set to 150 L/h to protect the cone from residue accumulation during operation. Both positive and negative ion modes (RPC+ and RPC-, respectively) were used. Raw spectra were converted into mzML files using MSConvert [6] and processed with XCMS 3.6.1 in R [7]. Peak picking and peak grouping were performed using in-house scripts in R and matrices were normalized using a median fold change approach. Log transformation, scaling and data analysis was performed in SIMCA 15.0 (Umetrics, Umea, Sweden).

#### Skin biopsy

The abdominal site was cleansed with (3) Chloraprep swabs (chlorhexidine 2% and isopropyl alcohol 70%, Becton Dickinson, Canaan, CT). Using sterile technique, local anaesthesia was obtained by infiltration of the area with 4mls of lidocaine 1% (Hospira, Inc., Lake Forest, IL) mixed with 1ml sodium bicarbonate. The skin biopsy was performed using a 6 mm punch (Miltex Instruments, York, PA). Fat tissue was carefully removed from the skin core of the biopsy using an 11-blade scalpel. The dermis and epidermis were divided into two halves, one half placed in a cryomold for OTC flash freezing (Agar Scientific, Essex, UK) and stored at -−80° C), and the other half was placed in RNAlater Stabilization Solution (Thermo Fisher Scientific, Fair Lawn, N.J.), refrigerated for 24 hours, and then frozen at −80° C. The fat tissue removed from the biopsy also was divided between RNAlater and a dry Sarstedt tube that was flash frozen in liquid nitrogen and placed in −80C. The adipose tissue processed in RNAlater was refrigerated for 24 hours and then frozen at −80C. The biopsy site was sutured closed and a dry sterile dressing was applied.

## Supplemental Tables

**Table S1.**
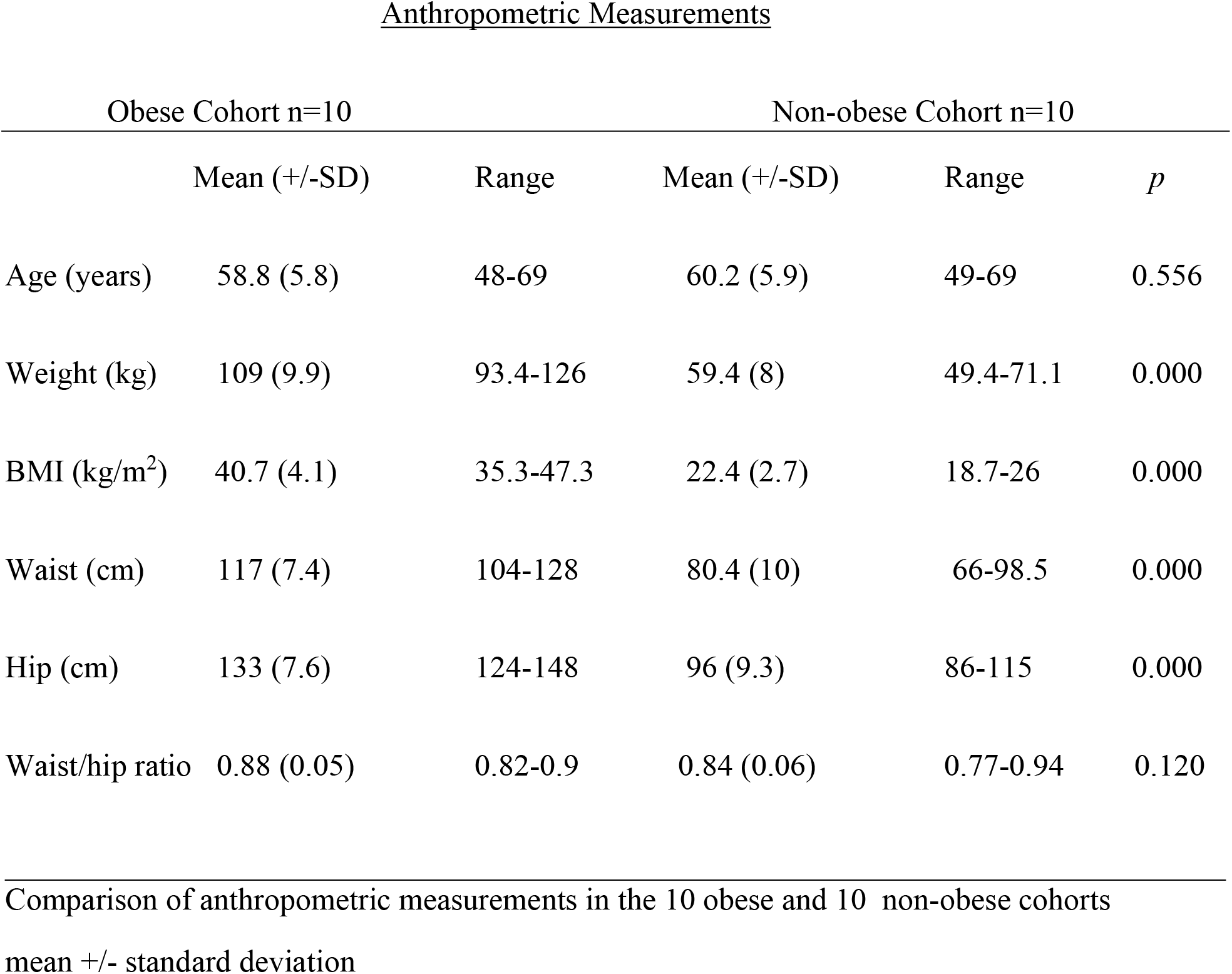
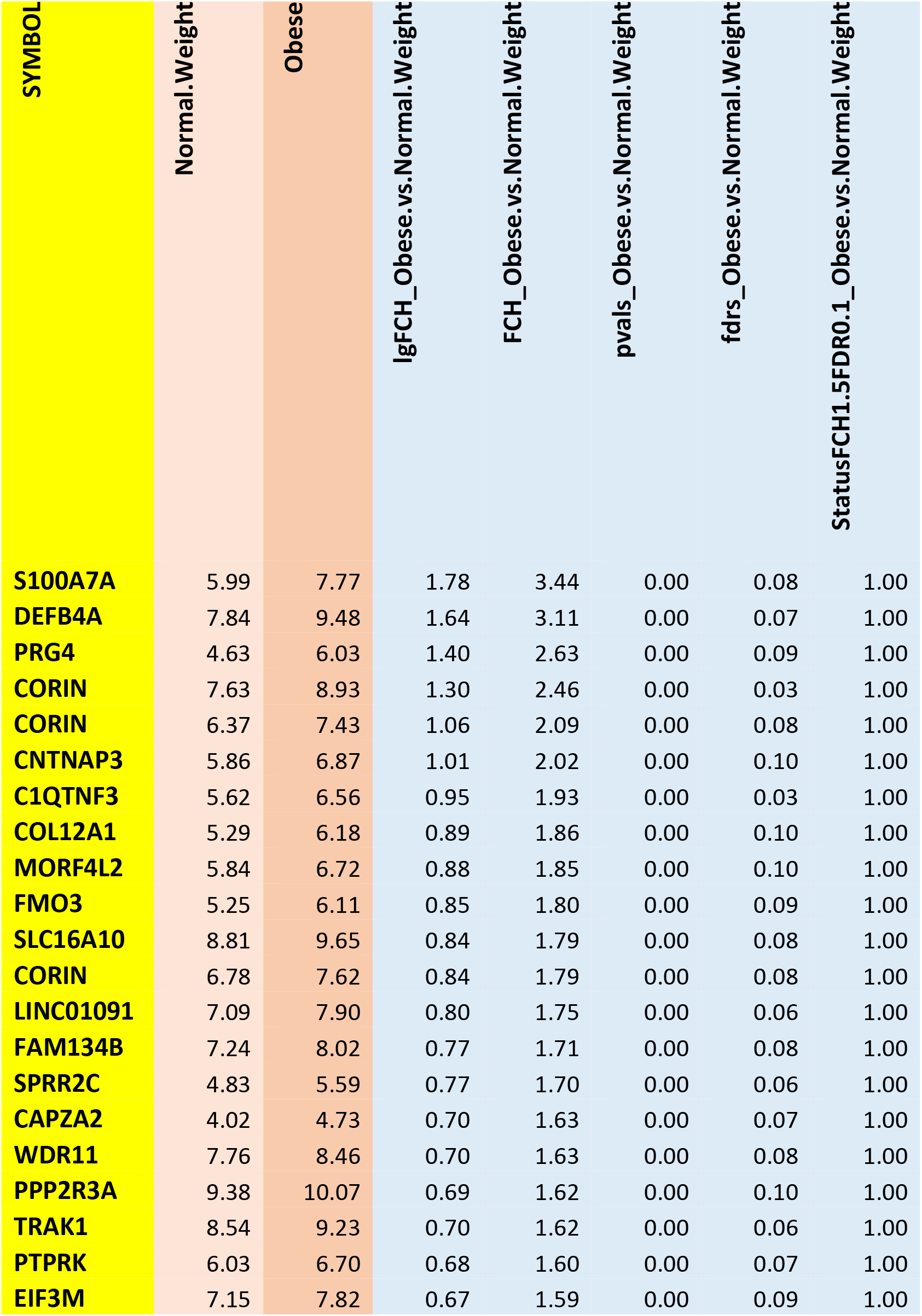
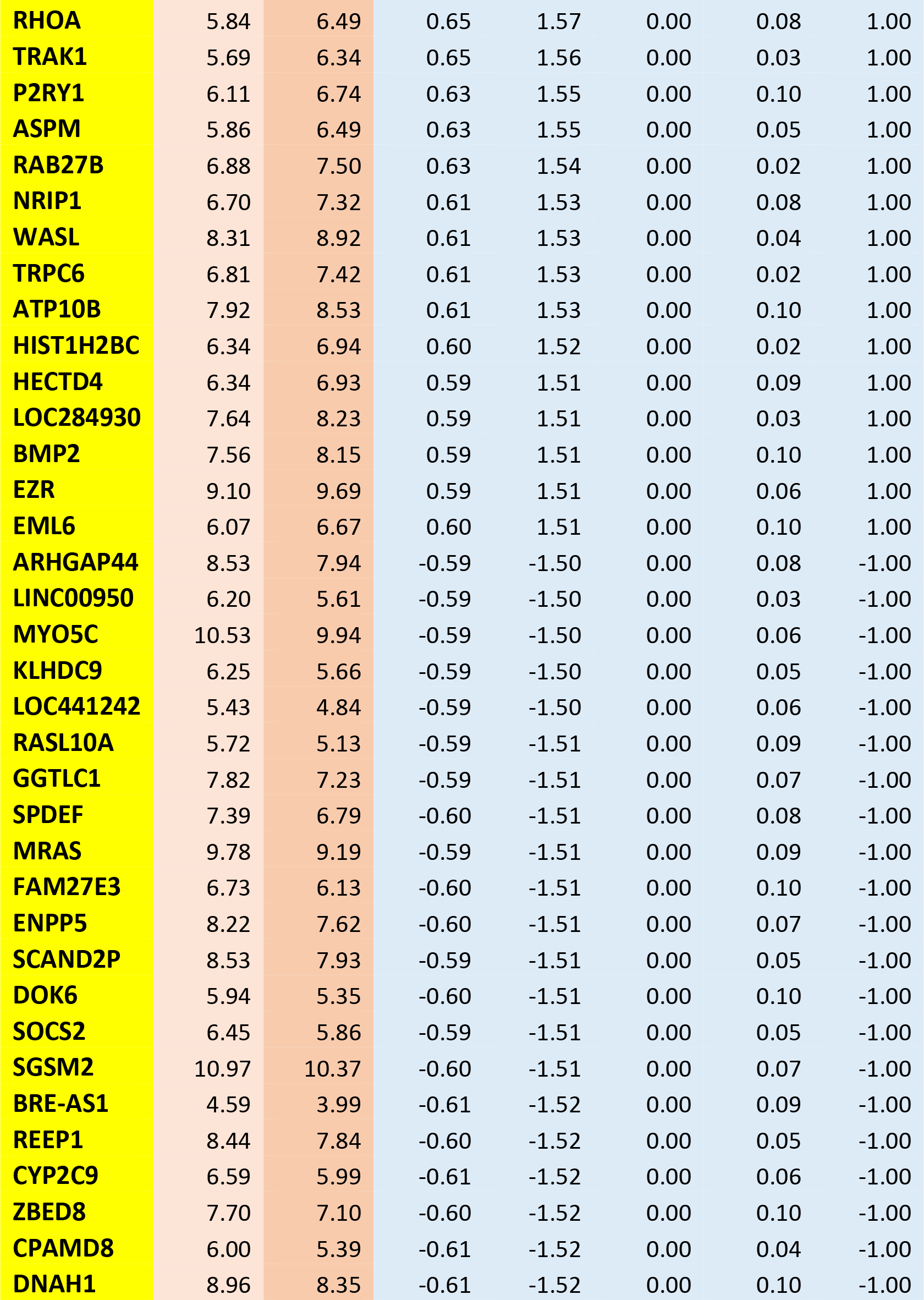
Skin: Obese vs Non-obese 1^st^ 70 genes

Anthropometric Measurements
|  | Obese Cohort n=10 |  | Non-obese Cohort n=10 |  | *p* |
| --- | --- | --- | --- | --- | --- |
|  | Mean (+/-SD) | Range | Mean (+/-SD) | Range |  |
| Age (years) | 58.8 (5.8) | 48-69 | 60.2 (5.9) | 49-69 | 0.556 |
| Weight (kg) | 109 (9.9) | 93.4-126 | 59.4 (8) | 49.4-71.1 | 0.000 |
| BMI (kg/m<sup>2</sup>) | 40.7 (4.1) | 35.3-47.3 | 22.4 (2.7) | 18.7-26 | 0.000 |
| Waist (cm) | 117 (7.4) | 104-128 | 80.4 (10) | 66-98.5 | 0.000 |
| Hip (cm) | 133 (7.6) | 124-148 | 96 (9.3) | 86-115 | 0.000 |
| Waist/hip ratio | 0.88 (0.05) | 0.82-0.9 | 0.84 (0.06) | 0.77-0.94 | 0.120 |

| SYMBOL | Normal.Weight | Obese | lgFCH_Obese.vs.Normal.Weight | FCH_Obese.vs.Normal.Weight | pvals_Obese.vs.Normal.Weight | fdrs_Obese.vs.Normal.Weight | StatusFCH1.5FDR0.1_Obese.vs.Normal.Weight |
| --- | --- | --- | --- | --- | --- | --- | --- |
| S100A7A | 5.99 | 7.77 | 1.78 | 3.44 | 0.00 | 0.08 | 1.00 |
| DEFB4A | 7.84 | 9.48 | 1.64 | 3.11 | 0.00 | 0.07 | 1.00 |
| PRG4 | 4.63 | 6.03 | 1.40 | 2.63 | 0.00 | 0.09 | 1.00 |
| CORIN | 7.63 | 8.93 | 1.30 | 2.46 | 0.00 | 0.03 | 1.00 |
| CORIN | 6.37 | 7.43 | 1.06 | 2.09 | 0.00 | 0.08 | 1.00 |
| CNTNAP3 | 5.86 | 6.87 | 1.01 | 2.02 | 0.00 | 0.10 | 1.00 |
| C1QTNF3 | 5.62 | 6.56 | 0.95 | 1.93 | 0.00 | 0.03 | 1.00 |
| COL12A1 | 5.29 | 6.18 | 0.89 | 1.86 | 0.00 | 0.10 | 1.00 |
| MORF4L2 | 5.84 | 6.72 | 0.88 | 1.85 | 0.00 | 0.10 | 1.00 |
| FMO3 | 5.25 | 6.11 | 0.85 | 1.80 | 0.00 | 0.09 | 1.00 |
| SLC16A10 | 8.81 | 9.65 | 0.84 | 1.79 | 0.00 | 0.08 | 1.00 |
| CORIN | 6.78 | 7.62 | 0.84 | 1.79 | 0.00 | 0.08 | 1.00 |
| LINC01091 | 7.09 | 7.90 | 0.80 | 1.75 | 0.00 | 0.06 | 1.00 |
| FAM134B | 7.24 | 8.02 | 0.77 | 1.71 | 0.00 | 0.08 | 1.00 |
| SPRR2C | 4.83 | 5.59 | 0.77 | 1.70 | 0.00 | 0.06 | 1.00 |
| CAPZA2 | 4.02 | 4.73 | 0.70 | 1.63 | 0.00 | 0.07 | 1.00 |
| WDR11 | 7.76 | 8.46 | 0.70 | 1.63 | 0.00 | 0.08 | 1.00 |
| PPP2R3A | 9.38 | 10.07 | 0.69 | 1.62 | 0.00 | 0.10 | 1.00 |
| TRAK1 | 8.54 | 9.23 | 0.70 | 1.62 | 0.00 | 0.06 | 1.00 |
| PTPRK | 6.03 | 6.70 | 0.68 | 1.60 | 0.00 | 0.07 | 1.00 |
| EIF3M | 7.15 | 7.82 | 0.67 | 1.59 | 0.00 | 0.09 | 1.00 |

|  |  |  |  |  |  |  |  |
| --- | --- | --- | --- | --- | --- | --- | --- |
| **RHOA** | 5.84 | 6.49 | 0.65 | 1.57 | 0.00 | 0.08 | 1.00 |
| **TRAK1** | 5.69 | 6.34 | 0.65 | 1.56 | 0.00 | 0.03 | 1.00 |
| **P2RY1** | 6.11 | 6.74 | 0.63 | 1.55 | 0.00 | 0.10 | 1.00 |
| **ASPM** | 5.86 | 6.49 | 0.63 | 1.55 | 0.00 | 0.05 | 1.00 |
| **RAB27B** | 6.88 | 7.50 | 0.63 | 1.54 | 0.00 | 0.02 | 1.00 |
| **NRIP1** | 6.70 | 7.32 | 0.61 | 1.53 | 0.00 | 0.08 | 1.00 |
| **WASL** | 8.31 | 8.92 | 0.61 | 1.53 | 0.00 | 0.04 | 1.00 |
| **TRPC6** | 6.81 | 7.42 | 0.61 | 1.53 | 0.00 | 0.02 | 1.00 |
| **ATP10B** | 7.92 | 8.53 | 0.61 | 1.53 | 0.00 | 0.10 | 1.00 |
| **HIST1H2BC** | 6.34 | 6.94 | 0.60 | 1.52 | 0.00 | 0.02 | 1.00 |
| **HECTD4** | 6.34 | 6.93 | 0.59 | 1.51 | 0.00 | 0.09 | 1.00 |
| **LOC284930** | 7.64 | 8.23 | 0.59 | 1.51 | 0.00 | 0.03 | 1.00 |
| **BMP2** | 7.56 | 8.15 | 0.59 | 1.51 | 0.00 | 0.10 | 1.00 |
| **EZR** | 9.10 | 9.69 | 0.59 | 1.51 | 0.00 | 0.06 | 1.00 |
| **EML6** | 6.07 | 6.67 | 0.60 | 1.51 | 0.00 | 0.10 | 1.00 |
| **ARHGAP44** | 8.53 | 7.94 | -0.59 | -1.50 | 0.00 | 0.08 | -1.00 |
| **LINC00950** | 6.20 | 5.61 | -0.59 | -1.50 | 0.00 | 0.03 | -1.00 |
| **MYO5C** | 10.53 | 9.94 | -0.59 | -1.50 | 0.00 | 0.06 | -1.00 |
| **KLHDC9** | 6.25 | 5.66 | -0.59 | -1.50 | 0.00 | 0.05 | -1.00 |
| **LOC441242** | 5.43 | 4.84 | -0.59 | -1.50 | 0.00 | 0.06 | -1.00 |
| **RASL10A** | 5.72 | 5.13 | -0.59 | -1.51 | 0.00 | 0.09 | -1.00 |
| **GGTLC1** | 7.82 | 7.23 | -0.59 | -1.51 | 0.00 | 0.07 | -1.00 |
| **SPDEF** | 7.39 | 6.79 | -0.60 | -1.51 | 0.00 | 0.08 | -1.00 |
| **MRAS** | 9.78 | 9.19 | -0.59 | -1.51 | 0.00 | 0.09 | -1.00 |
| **FAM27E3** | 6.73 | 6.13 | -0.60 | -1.51 | 0.00 | 0.10 | -1.00 |
| **ENPP5** | 8.22 | 7.62 | -0.60 | -1.51 | 0.00 | 0.07 | -1.00 |
| **SCAND2P** | 8.53 | 7.93 | -0.59 | -1.51 | 0.00 | 0.05 | -1.00 |
| **DOK6** | 5.94 | 5.35 | -0.60 | -1.51 | 0.00 | 0.10 | -1.00 |
| **SOCS2** | 6.45 | 5.86 | -0.59 | -1.51 | 0.00 | 0.05 | -1.00 |
| **SGSM2** | 10.97 | 10.37 | -0.60 | -1.51 | 0.00 | 0.07 | -1.00 |
| **BRE-AS1** | 4.59 | 3.99 | -0.61 | -1.52 | 0.00 | 0.09 | -1.00 |
| **REEP1** | 8.44 | 7.84 | -0.60 | -1.52 | 0.00 | 0.05 | -1.00 |
| **CYP2C9** | 6.59 | 5.99 | -0.61 | -1.52 | 0.00 | 0.06 | -1.00 |
| **ZBED8** | 7.70 | 7.10 | -0.60 | -1.52 | 0.00 | 0.10 | -1.00 |
| **CPAMD8** | 6.00 | 5.39 | -0.61 | -1.52 | 0.00 | 0.04 | -1.00 |
| **DNAH1** | 8.96 | 8.35 | -0.61 | -1.52 | 0.00 | 0.10 | -1.00 |

**Table S2.**
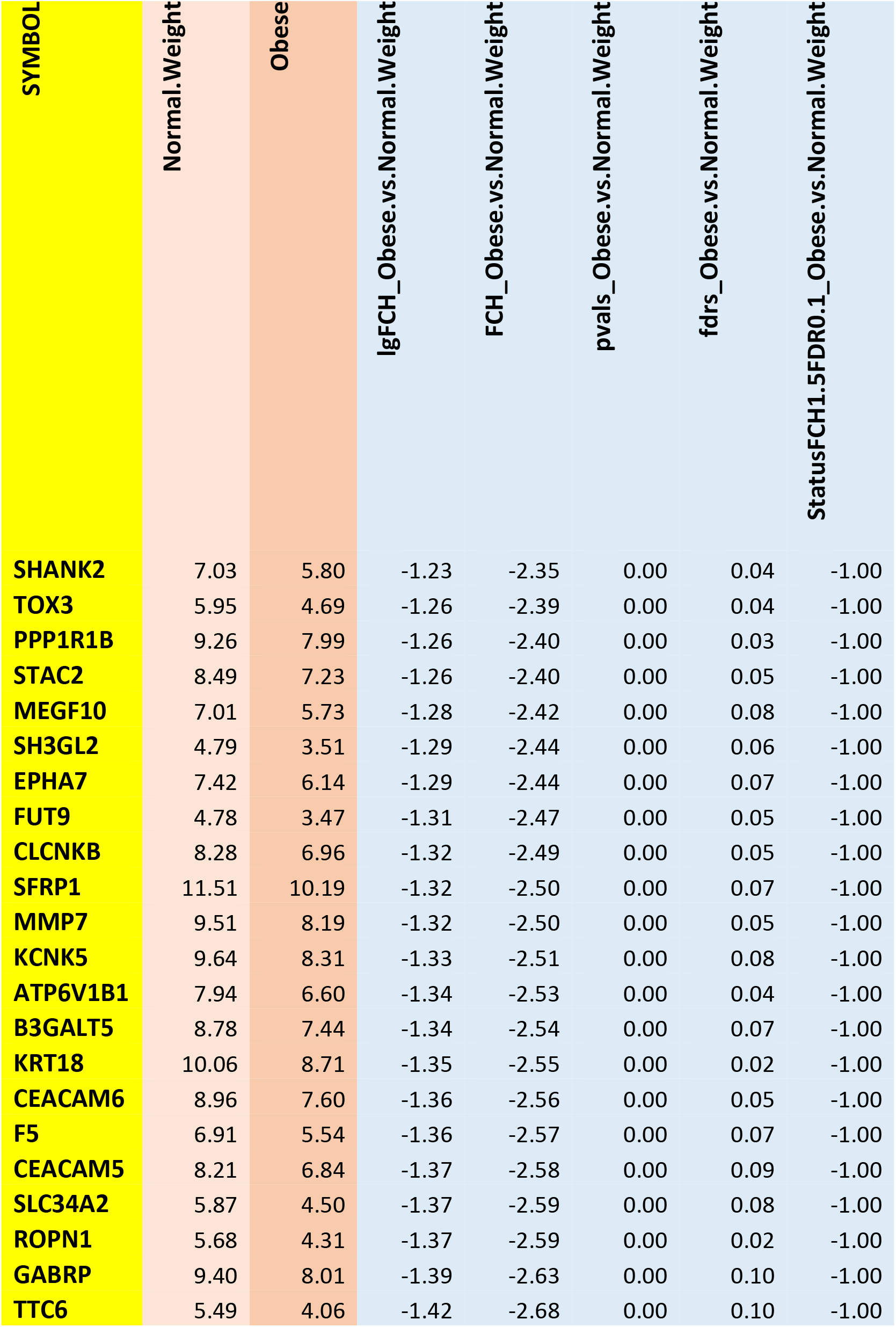
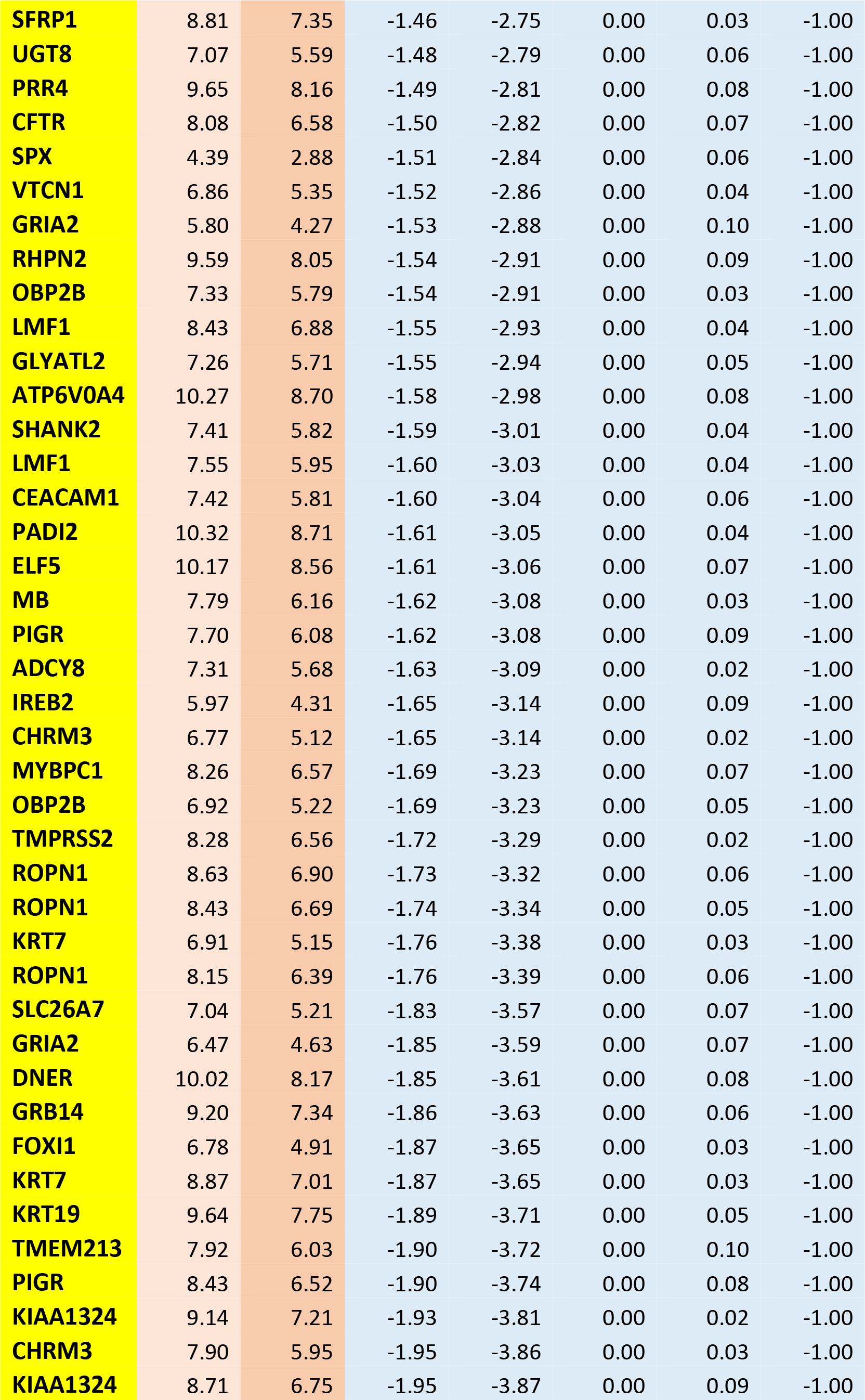
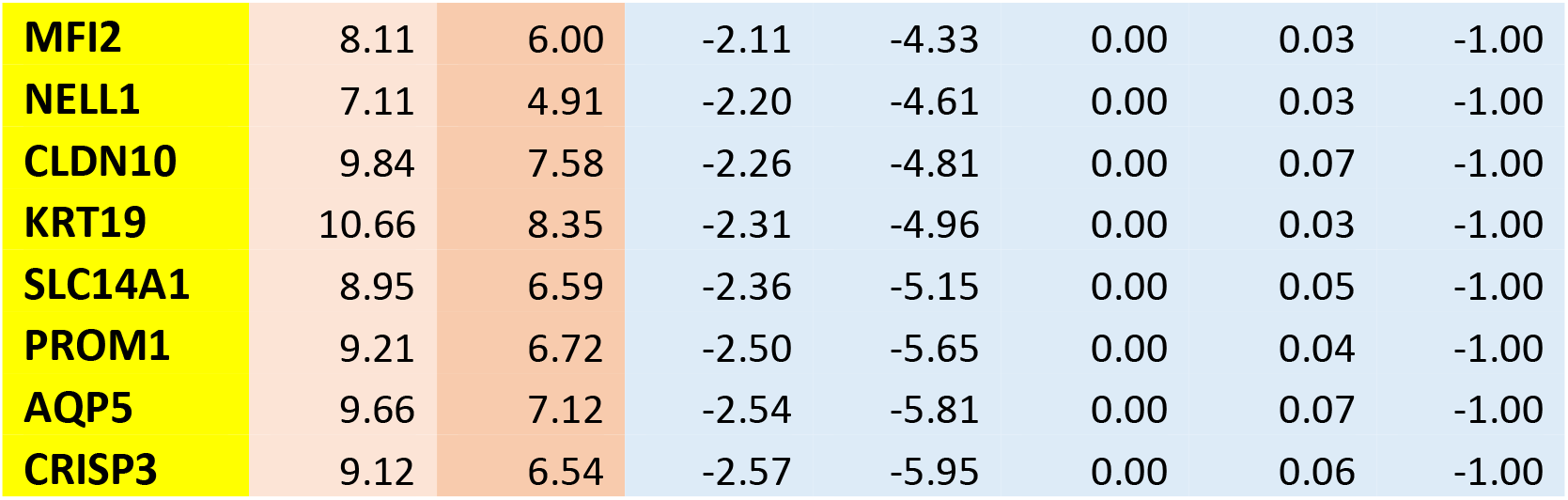
Skin: Obese vs Non-obese last 70 genes

Skin: Obese vs Non-obese last 70 genes
| SYMBOL | Normal.Weight | Obese | lgFCH_Obese.vs.Normal.Weight | FCH_Obese.vs.Normal.Weight | pvals_Obese.vs.Normal.Weight | fdrs_Obese.vs.Normal.Weight | StatusFCH1.5FDR0.1_Obese.vs.Normal.Weight |
| --- | --- | --- | --- | --- | --- | --- | --- |
| SHANK2 | 7.03 | 5.80 | -1.23 | -2.35 | 0.00 | 0.04 | -1.00 |
| TOX3 | 5.95 | 4.69 | -1.26 | -2.39 | 0.00 | 0.04 | -1.00 |
| PPP1R1B | 9.26 | 7.99 | -1.26 | -2.40 | 0.00 | 0.03 | -1.00 |
| STAC2 | 8.49 | 7.23 | -1.26 | -2.40 | 0.00 | 0.05 | -1.00 |
| MEGF10 | 7.01 | 5.73 | -1.28 | -2.42 | 0.00 | 0.08 | -1.00 |
| SH3GL2 | 4.79 | 3.51 | -1.29 | -2.44 | 0.00 | 0.06 | -1.00 |
| EPHA7 | 7.42 | 6.14 | -1.29 | -2.44 | 0.00 | 0.07 | -1.00 |
| FUT9 | 4.78 | 3.47 | -1.31 | -2.47 | 0.00 | 0.05 | -1.00 |
| CLCNKB | 8.28 | 6.96 | -1.32 | -2.49 | 0.00 | 0.05 | -1.00 |
| SFRP1 | 11.51 | 10.19 | -1.32 | -2.50 | 0.00 | 0.07 | -1.00 |
| MMP7 | 9.51 | 8.19 | -1.32 | -2.50 | 0.00 | 0.05 | -1.00 |
| KCNK5 | 9.64 | 8.31 | -1.33 | -2.51 | 0.00 | 0.08 | -1.00 |
| ATP6V1B1 | 7.94 | 6.60 | -1.34 | -2.53 | 0.00 | 0.04 | -1.00 |
| B3GALT5 | 8.78 | 7.44 | -1.34 | -2.54 | 0.00 | 0.07 | -1.00 |
| KRT18 | 10.06 | 8.71 | -1.35 | -2.55 | 0.00 | 0.02 | -1.00 |
| CEACAM6 | 8.96 | 7.60 | -1.36 | -2.56 | 0.00 | 0.05 | -1.00 |
| F5 | 6.91 | 5.54 | -1.36 | -2.57 | 0.00 | 0.07 | -1.00 |
| CEACAM5 | 8.21 | 6.84 | -1.37 | -2.58 | 0.00 | 0.09 | -1.00 |
| SLC34A2 | 5.87 | 4.50 | -1.37 | -2.59 | 0.00 | 0.08 | -1.00 |
| ROPN1 | 5.68 | 4.31 | -1.37 | -2.59 | 0.00 | 0.02 | -1.00 |
| GABRP | 9.40 | 8.01 | -1.39 | -2.63 | 0.00 | 0.10 | -1.00 |
| TTC6 | 5.49 | 4.06 | -1.42 | -2.68 | 0.00 | 0.10 | -1.00 |
| **SFRP1** | 8.81 | 7.35 | -1.46 | -2.75 | 0.00 | 0.03 | -1.00 |
| **UGT8** | 7.07 | 5.59 | -1.48 | -2.79 | 0.00 | 0.06 | -1.00 |
| **PRR4** | 9.65 | 8.16 | -1.49 | -2.81 | 0.00 | 0.08 | -1.00 |
| **CFTR** | 8.08 | 6.58 | -1.50 | -2.82 | 0.00 | 0.07 | -1.00 |
| **SPX** | 4.39 | 2.88 | -1.51 | -2.84 | 0.00 | 0.06 | -1.00 |
| **VTCN1** | 6.86 | 5.35 | -1.52 | -2.86 | 0.00 | 0.04 | -1.00 |
| **GRIA2** | 5.80 | 4.27 | -1.53 | -2.88 | 0.00 | 0.10 | -1.00 |
| **RHPN2** | 9.59 | 8.05 | -1.54 | -2.91 | 0.00 | 0.09 | -1.00 |
| **OBP2B** | 7.33 | 5.79 | -1.54 | -2.91 | 0.00 | 0.03 | -1.00 |
| **LMF1** | 8.43 | 6.88 | -1.55 | -2.93 | 0.00 | 0.04 | -1.00 |
| **GLYATL2** | 7.26 | 5.71 | -1.55 | -2.94 | 0.00 | 0.05 | -1.00 |
| **ATP6V0A4** | 10.27 | 8.70 | -1.58 | -2.98 | 0.00 | 0.08 | -1.00 |
| **SHANK2** | 7.41 | 5.82 | -1.59 | -3.01 | 0.00 | 0.04 | -1.00 |
| **LMF1** | 7.55 | 5.95 | -1.60 | -3.03 | 0.00 | 0.04 | -1.00 |
| **CEACAM1** | 7.42 | 5.81 | -1.60 | -3.04 | 0.00 | 0.06 | -1.00 |
| **PADI2** | 10.32 | 8.71 | -1.61 | -3.05 | 0.00 | 0.04 | -1.00 |
| **ELF5** | 10.17 | 8.56 | -1.61 | -3.06 | 0.00 | 0.07 | -1.00 |
| **MB** | 7.79 | 6.16 | -1.62 | -3.08 | 0.00 | 0.03 | -1.00 |
| **PIGR** | 7.70 | 6.08 | -1.62 | -3.08 | 0.00 | 0.09 | -1.00 |
| **ADCY8** | 7.31 | 5.68 | -1.63 | -3.09 | 0.00 | 0.02 | -1.00 |
| **IREB2** | 5.97 | 4.31 | -1.65 | -3.14 | 0.00 | 0.09 | -1.00 |
| **CHRM3** | 6.77 | 5.12 | -1.65 | -3.14 | 0.00 | 0.02 | -1.00 |
| **MYBPC1** | 8.26 | 6.57 | -1.69 | -3.23 | 0.00 | 0.07 | -1.00 |
| **OBP2B** | 6.92 | 5.22 | -1.69 | -3.23 | 0.00 | 0.05 | -1.00 |
| **TMPRSS2** | 8.28 | 6.56 | -1.72 | -3.29 | 0.00 | 0.02 | -1.00 |
| **ROPN1** | 8.63 | 6.90 | -1.73 | -3.32 | 0.00 | 0.06 | -1.00 |
| **ROPN1** | 8.43 | 6.69 | -1.74 | -3.34 | 0.00 | 0.05 | -1.00 |
| **KRT7** | 6.91 | 5.15 | -1.76 | -3.38 | 0.00 | 0.03 | -1.00 |
| **ROPN1** | 8.15 | 6.39 | -1.76 | -3.39 | 0.00 | 0.06 | -1.00 |
| **SLC26A7** | 7.04 | 5.21 | -1.83 | -3.57 | 0.00 | 0.07 | -1.00 |
| **GRIA2** | 6.47 | 4.63 | -1.85 | -3.59 | 0.00 | 0.07 | -1.00 |
| **DNER** | 10.02 | 8.17 | -1.85 | -3.61 | 0.00 | 0.08 | -1.00 |
| **GRB14** | 9.20 | 7.34 | -1.86 | -3.63 | 0.00 | 0.06 | -1.00 |
| **FOXI1** | 6.78 | 4.91 | -1.87 | -3.65 | 0.00 | 0.03 | -1.00 |
| **KRT7** | 8.87 | 7.01 | -1.87 | -3.65 | 0.00 | 0.03 | -1.00 |
| **KRT19** | 9.64 | 7.75 | -1.89 | -3.71 | 0.00 | 0.05 | -1.00 |
| **TMEM213** | 7.92 | 6.03 | -1.90 | -3.72 | 0.00 | 0.10 | -1.00 |
| **PIGR** | 8.43 | 6.52 | -1.90 | -3.74 | 0.00 | 0.08 | -1.00 |
| **KIAA1324** | 9.14 | 7.21 | -1.93 | -3.81 | 0.00 | 0.02 | -1.00 |
| **CHRM3** | 7.90 | 5.95 | -1.95 | -3.86 | 0.00 | 0.03 | -1.00 |
| **KIAA1324** | 8.71 | 6.75 | -1.95 | -3.87 | 0.00 | 0.09 | -1.00 |
| **MFI2** | 8.11 | 6.00 | -2.11 | -4.33 | 0.00 | 0.03 | -1.00 |
| **NELL1** | 7.11 | 4.91 | -2.20 | -4.61 | 0.00 | 0.03 | -1.00 |
| **CLDN10** | 9.84 | 7.58 | -2.26 | -4.81 | 0.00 | 0.07 | -1.00 |
| **KRT19** | 10.66 | 8.35 | -2.31 | -4.96 | 0.00 | 0.03 | -1.00 |
| **SLC14A1** | 8.95 | 6.59 | -2.36 | -5.15 | 0.00 | 0.05 | -1.00 |
| **PROM1** | 9.21 | 6.72 | -2.50 | -5.65 | 0.00 | 0.04 | -1.00 |
| **AQP5** | 9.66 | 7.12 | -2.54 | -5.81 | 0.00 | 0.07 | -1.00 |
| **CRISP3** | 9.12 | 6.54 | -2.57 | -5.95 | 0.00 | 0.06 | -1.00 |

**Table S3.**
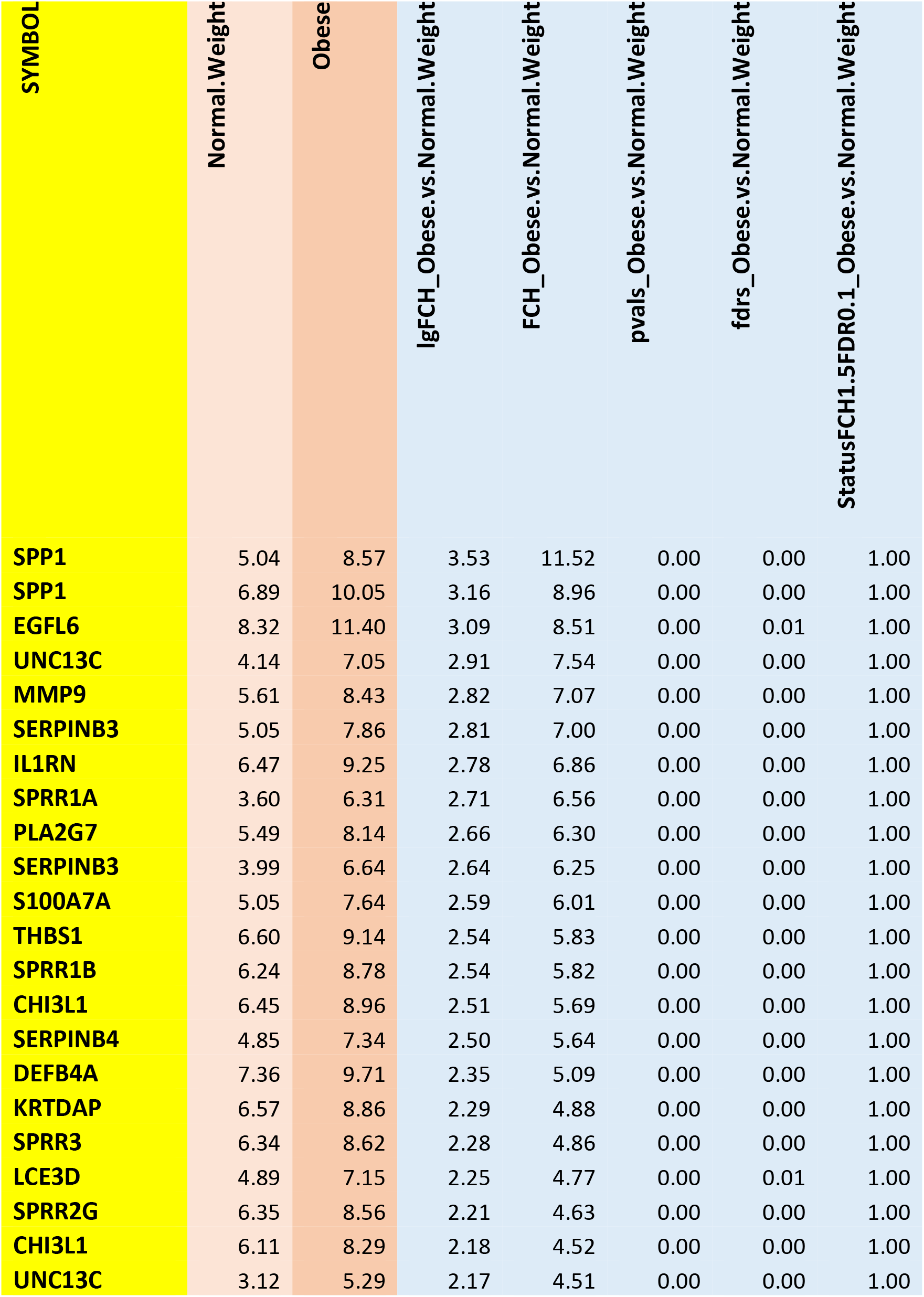
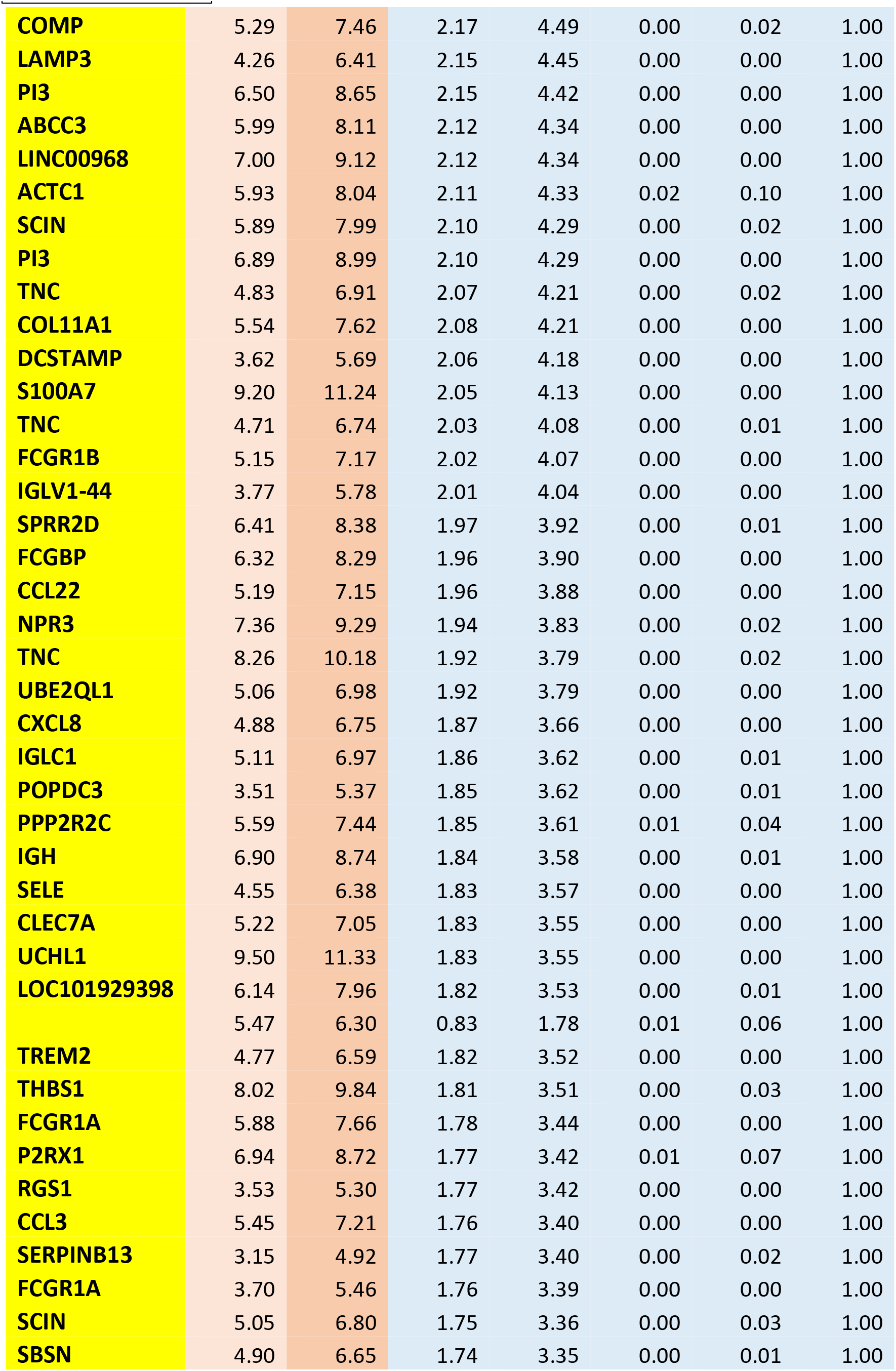
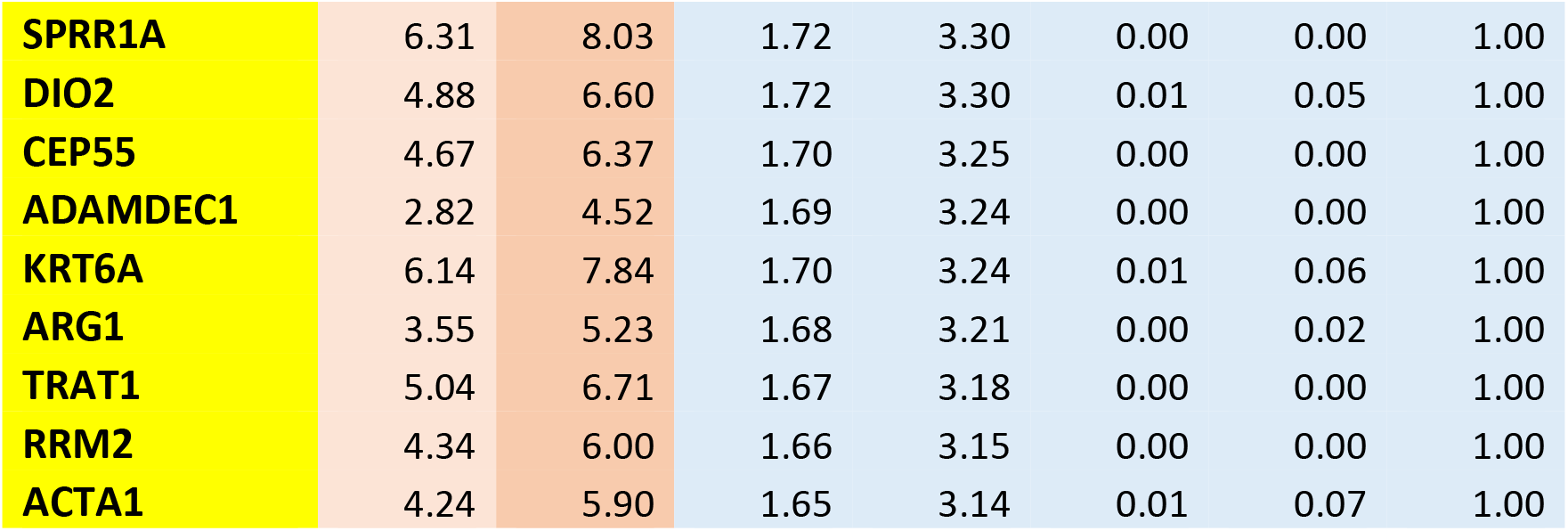
Fat: Obese vs Non-obese 1^st^ 70 genes

| SYMBOL | Normal.Weight | Obese | lgFCH_Obese.vs.Normal.Weight | FCH_Obese.vs.Normal.Weight | pvals_Obese.vs.Normal.Weight | fdrs_Obese.vs.Normal.Weight | StatusFCH1.5FDR0.1_Obese.vs.Normal.Weight |
| --- | --- | --- | --- | --- | --- | --- | --- |
| SPP1 | 5.04 | 8.57 | 3.53 | 11.52 | 0.00 | 0.00 | 1.00 |
| SPP1 | 6.89 | 10.05 | 3.16 | 8.96 | 0.00 | 0.00 | 1.00 |
| EGFL6 | 8.32 | 11.40 | 3.09 | 8.51 | 0.00 | 0.01 | 1.00 |
| UNC13C | 4.14 | 7.05 | 2.91 | 7.54 | 0.00 | 0.00 | 1.00 |
| MMP9 | 5.61 | 8.43 | 2.82 | 7.07 | 0.00 | 0.00 | 1.00 |
| SERPINB3 | 5.05 | 7.86 | 2.81 | 7.00 | 0.00 | 0.00 | 1.00 |
| IL1RN | 6.47 | 9.25 | 2.78 | 6.86 | 0.00 | 0.00 | 1.00 |
| SPRR1A | 3.60 | 6.31 | 2.71 | 6.56 | 0.00 | 0.00 | 1.00 |
| PLA2G7 | 5.49 | 8.14 | 2.66 | 6.30 | 0.00 | 0.00 | 1.00 |
| SERPINB3 | 3.99 | 6.64 | 2.64 | 6.25 | 0.00 | 0.00 | 1.00 |
| S100A7A | 5.05 | 7.64 | 2.59 | 6.01 | 0.00 | 0.00 | 1.00 |
| THBS1 | 6.60 | 9.14 | 2.54 | 5.83 | 0.00 | 0.00 | 1.00 |
| SPRR1B | 6.24 | 8.78 | 2.54 | 5.82 | 0.00 | 0.00 | 1.00 |
| CHI3L1 | 6.45 | 8.96 | 2.51 | 5.69 | 0.00 | 0.00 | 1.00 |
| SERPINB4 | 4.85 | 7.34 | 2.50 | 5.64 | 0.00 | 0.00 | 1.00 |
| DEFB4A | 7.36 | 9.71 | 2.35 | 5.09 | 0.00 | 0.00 | 1.00 |
| KRTDAP | 6.57 | 8.86 | 2.29 | 4.88 | 0.00 | 0.00 | 1.00 |
| SPRR3 | 6.34 | 8.62 | 2.28 | 4.86 | 0.00 | 0.00 | 1.00 |
| LCE3D | 4.89 | 7.15 | 2.25 | 4.77 | 0.00 | 0.01 | 1.00 |
| SPRR2G | 6.35 | 8.56 | 2.21 | 4.63 | 0.00 | 0.00 | 1.00 |
| CHI3L1 | 6.11 | 8.29 | 2.18 | 4.52 | 0.00 | 0.00 | 1.00 |
| UNC13C | 3.12 | 5.29 | 2.17 | 4.51 | 0.00 | 0.00 | 1.00 |
| COMP | 5.29 | 7.46 | 2.17 | 4.49 | 0.00 | 0.02 | 1.00 |
| LAMP3 | 4.26 | 6.41 | 2.15 | 4.45 | 0.00 | 0.00 | 1.00 |
| PI3 | 6.50 | 8.65 | 2.15 | 4.42 | 0.00 | 0.00 | 1.00 |
| ABCC3 | 5.99 | 8.11 | 2.12 | 4.34 | 0.00 | 0.00 | 1.00 |
| LINC00968 | 7.00 | 9.12 | 2.12 | 4.34 | 0.00 | 0.00 | 1.00 |
| ACTC1 | 5.93 | 8.04 | 2.11 | 4.33 | 0.02 | 0.10 | 1.00 |
| SCIN | 5.89 | 7.99 | 2.10 | 4.29 | 0.00 | 0.02 | 1.00 |
| PI3 | 6.89 | 8.99 | 2.10 | 4.29 | 0.00 | 0.00 | 1.00 |
| TNC | 4.83 | 6.91 | 2.07 | 4.21 | 0.00 | 0.02 | 1.00 |
| COL11A1 | 5.54 | 7.62 | 2.08 | 4.21 | 0.00 | 0.00 | 1.00 |
| DCSTAMP | 3.62 | 5.69 | 2.06 | 4.18 | 0.00 | 0.00 | 1.00 |
| S100A7 | 9.20 | 11.24 | 2.05 | 4.13 | 0.00 | 0.00 | 1.00 |
| TNC | 4.71 | 6.74 | 2.03 | 4.08 | 0.00 | 0.01 | 1.00 |
| FCGR1B | 5.15 | 7.17 | 2.02 | 4.07 | 0.00 | 0.00 | 1.00 |
| IGLV1-44 | 3.77 | 5.78 | 2.01 | 4.04 | 0.00 | 0.00 | 1.00 |
| SPRR2D | 6.41 | 8.38 | 1.97 | 3.92 | 0.00 | 0.01 | 1.00 |
| FCGBP | 6.32 | 8.29 | 1.96 | 3.90 | 0.00 | 0.00 | 1.00 |
| CCL22 | 5.19 | 7.15 | 1.96 | 3.88 | 0.00 | 0.00 | 1.00 |
| NPR3 | 7.36 | 9.29 | 1.94 | 3.83 | 0.00 | 0.02 | 1.00 |
| TNC | 8.26 | 10.18 | 1.92 | 3.79 | 0.00 | 0.02 | 1.00 |
| UBE2QL1 | 5.06 | 6.98 | 1.92 | 3.79 | 0.00 | 0.00 | 1.00 |
| CXCL8 | 4.88 | 6.75 | 1.87 | 3.66 | 0.00 | 0.00 | 1.00 |
| IGLC1 | 5.11 | 6.97 | 1.86 | 3.62 | 0.00 | 0.01 | 1.00 |
| POPDC3 | 3.51 | 5.37 | 1.85 | 3.62 | 0.00 | 0.01 | 1.00 |
| PPP2R2C | 5.59 | 7.44 | 1.85 | 3.61 | 0.01 | 0.04 | 1.00 |
| IGH | 6.90 | 8.74 | 1.84 | 3.58 | 0.00 | 0.01 | 1.00 |
| SELE | 4.55 | 6.38 | 1.83 | 3.57 | 0.00 | 0.00 | 1.00 |
| CLEC7A | 5.22 | 7.05 | 1.83 | 3.55 | 0.00 | 0.00 | 1.00 |
| UCHL1 | 9.50 | 11.33 | 1.83 | 3.55 | 0.00 | 0.00 | 1.00 |
| LOC101929398 | 6.14 | 7.96 | 1.82 | 3.53 | 0.00 | 0.01 | 1.00 |
|  | 5.47 | 6.30 | 0.83 | 1.78 | 0.01 | 0.06 | 1.00 |
| TREM2 | 4.77 | 6.59 | 1.82 | 3.52 | 0.00 | 0.00 | 1.00 |
| THBS1 | 8.02 | 9.84 | 1.81 | 3.51 | 0.00 | 0.03 | 1.00 |
| FCGR1A | 5.88 | 7.66 | 1.78 | 3.44 | 0.00 | 0.00 | 1.00 |
| P2RX1 | 6.94 | 8.72 | 1.77 | 3.42 | 0.01 | 0.07 | 1.00 |
| RGS1 | 3.53 | 5.30 | 1.77 | 3.42 | 0.00 | 0.00 | 1.00 |
| CCL3 | 5.45 | 7.21 | 1.76 | 3.40 | 0.00 | 0.00 | 1.00 |
| SERPINB13 | 3.15 | 4.92 | 1.77 | 3.40 | 0.00 | 0.02 | 1.00 |
| FCGR1A | 3.70 | 5.46 | 1.76 | 3.39 | 0.00 | 0.00 | 1.00 |
| SCIN | 5.05 | 6.80 | 1.75 | 3.36 | 0.00 | 0.03 | 1.00 |
| SBSN | 4.90 | 6.65 | 1.74 | 3.35 | 0.00 | 0.01 | 1.00 |
| **SPRR1A** | 6.31 | 8.03 | 1.72 | 3.30 | 0.00 | 0.00 | 1.00 |
| **DIO2** | 4.88 | 6.60 | 1.72 | 3.30 | 0.01 | 0.05 | 1.00 |
| **CEP55** | 4.67 | 6.37 | 1.70 | 3.25 | 0.00 | 0.00 | 1.00 |
| **ADAMDEC1** | 2.82 | 4.52 | 1.69 | 3.24 | 0.00 | 0.00 | 1.00 |
| **KRT6A** | 6.14 | 7.84 | 1.70 | 3.24 | 0.01 | 0.06 | 1.00 |
| **ARG1** | 3.55 | 5.23 | 1.68 | 3.21 | 0.00 | 0.02 | 1.00 |
| **TRAT1** | 5.04 | 6.71 | 1.67 | 3.18 | 0.00 | 0.00 | 1.00 |
| **RRM2** | 4.34 | 6.00 | 1.66 | 3.15 | 0.00 | 0.00 | 1.00 |
| **ACTA1** | 4.24 | 5.90 | 1.65 | 3.14 | 0.01 | 0.07 | 1.00 |

**Table S4.**
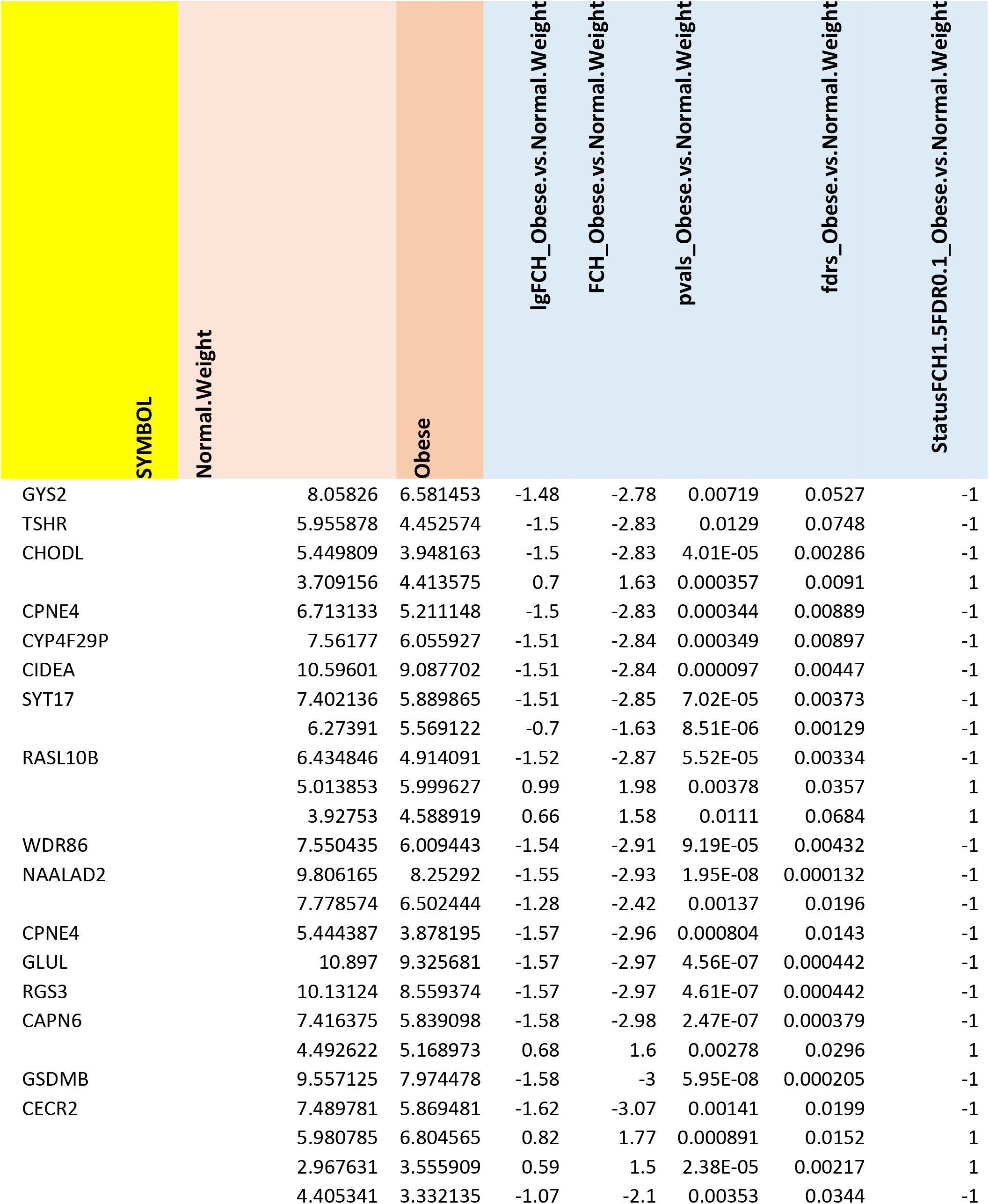
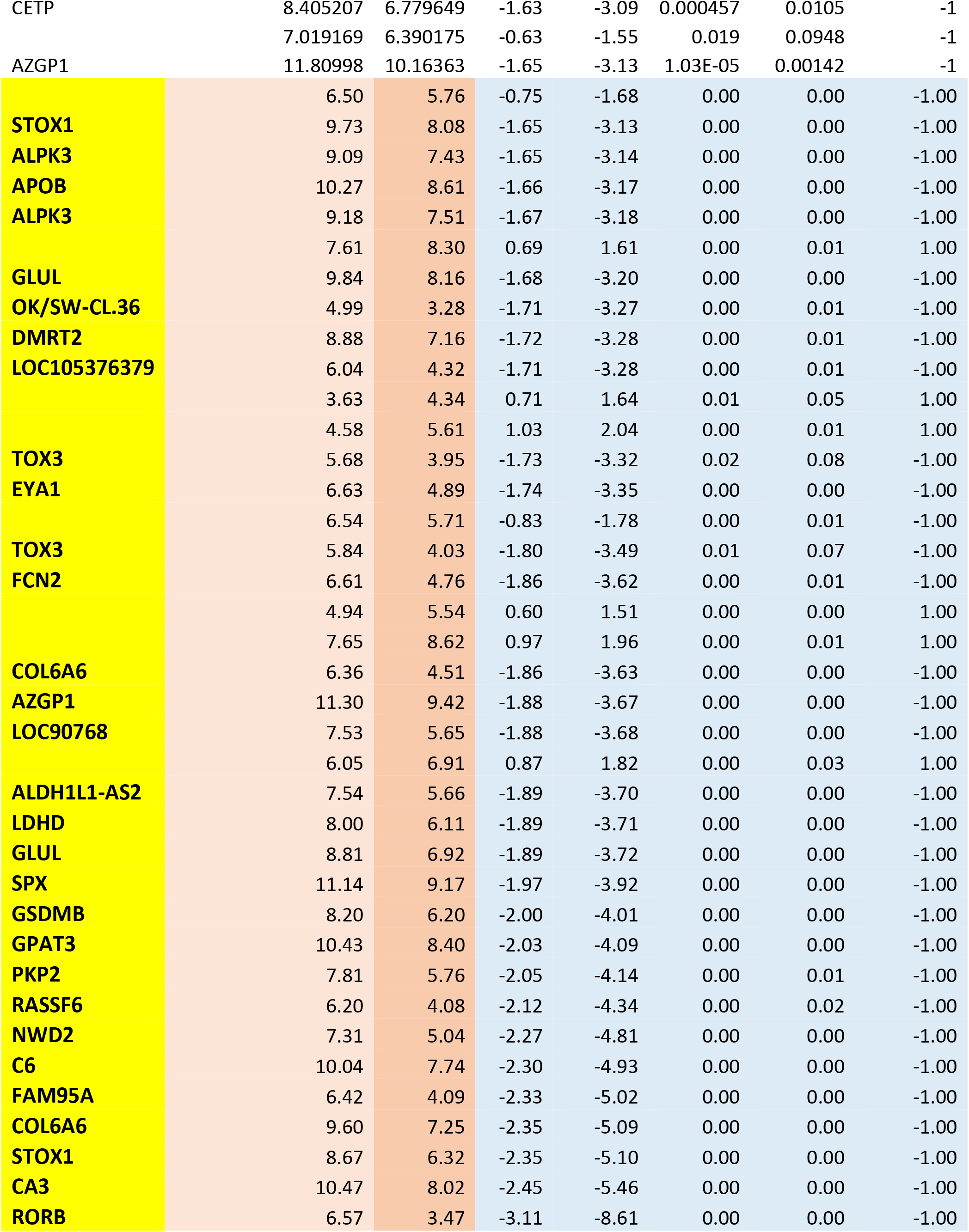
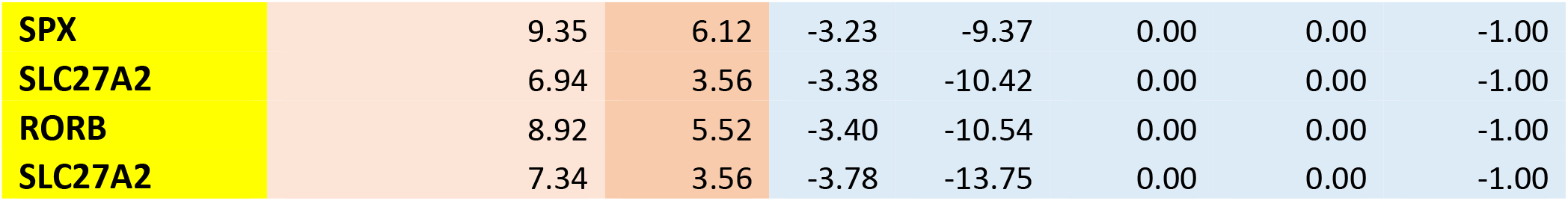
Fat: Obese vs Non-obese last 70 genes

Fat: Obese vs Non-obese last 70 genes
| SYMBOL | Normal.Weight | Obese | lgFCH_Obese.vs.Normal.Weight | FCH_Obese.vs.Normal.Weight | pvals_Obese.vs.Normal.Weight | fdrs_Obese.vs.Normal.Weight | StatusFCH1.5FDR0.1_Obese.vs.Normal.Weight |
| --- | --- | --- | --- | --- | --- | --- | --- |
| GYS2 | 8.05826 | 6.581453 | -1.48 | -2.78 | 0.00719 | 0.0527 | -1 |
| TSHR | 5.955878 | 4.452574 | -1.5 | -2.83 | 0.0129 | 0.0748 | -1 |
| CHODL | 5.449809 | 3.948163 | -1.5 | -2.83 | 4.01E-05 | 0.00286 | -1 |
|  | 3.709156 | 4.413575 | 0.7 | 1.63 | 0.000357 | 0.0091 | 1 |
| CPNE4 | 6.713133 | 5.211148 | -1.5 | -2.83 | 0.000344 | 0.00889 | -1 |
| CYP4F29P | 7.56177 | 6.055927 | -1.51 | -2.84 | 0.000349 | 0.00897 | -1 |
| CIDEA | 10.59601 | 9.087702 | -1.51 | -2.84 | 0.000097 | 0.00447 | -1 |
| SYT17 | 7.402136 | 5.889865 | -1.51 | -2.85 | 7.02E-05 | 0.00373 | -1 |
|  | 6.27391 | 5.569122 | -0.7 | -1.63 | 8.51E-06 | 0.00129 | -1 |
| RASL10B | 6.434846 | 4.914091 | -1.52 | -2.87 | 5.52E-05 | 0.00334 | -1 |
|  | 5.013853 | 5.999627 | 0.99 | 1.98 | 0.00378 | 0.0357 | 1 |
|  | 3.92753 | 4.588919 | 0.66 | 1.58 | 0.0111 | 0.0684 | 1 |
| WDR86 | 7.550435 | 6.009443 | -1.54 | -2.91 | 9.19E-05 | 0.00432 | -1 |
| NAALAD2 | 9.806165 | 8.25292 | -1.55 | -2.93 | 1.95E-08 | 0.000132 | -1 |
|  | 7.778574 | 6.502444 | -1.28 | -2.42 | 0.00137 | 0.0196 | -1 |
| CPNE4 | 5.444387 | 3.878195 | -1.57 | -2.96 | 0.000804 | 0.0143 | -1 |
| GLUL | 10.897 | 9.325681 | -1.57 | -2.97 | 4.56E-07 | 0.000442 | -1 |
| RGS3 | 10.13124 | 8.559374 | -1.57 | -2.97 | 4.61E-07 | 0.000442 | -1 |
| CAPN6 | 7.416375 | 5.839098 | -1.58 | -2.98 | 2.47E-07 | 0.000379 | -1 |
|  | 4.492622 | 5.168973 | 0.68 | 1.6 | 0.00278 | 0.0296 | 1 |
| GSDMB | 9.557125 | 7.974478 | -1.58 | -3 | 5.95E-08 | 0.000205 | -1 |
| CECR2 | 7.489781 | 5.869481 | -1.62 | -3.07 | 0.00141 | 0.0199 | -1 |
|  | 5.980785 | 6.804565 | 0.82 | 1.77 | 0.000891 | 0.0152 | 1 |
|  | 2.967631 | 3.555909 | 0.59 | 1.5 | 2.38E-05 | 0.00217 | 1 |
|  | 4.405341 | 3.332135 | -1.07 | -2.1 | 0.00353 | 0.0344 | -1 |
| CETP | 8.405207 | 6.779649 | -1.63 | -3.09 | 0.000457 | 0.0105 | -1 |
|  | 7.019169 | 6.390175 | -0.63 | -1.55 | 0.019 | 0.0948 | -1 |
| AZGP1 | 11.80998 | 10.16363 | -1.65 | -3.13 | 1.03E-05 | 0.00142 | -1 |
|  | 6.50 | 5.76 | -0.75 | -1.68 | 0.00 | 0.00 | -1.00 |
| **STOX1** | 9.73 | 8.08 | -1.65 | -3.13 | 0.00 | 0.00 | -1.00 |
| **ALPK3** | 9.09 | 7.43 | -1.65 | -3.14 | 0.00 | 0.00 | -1.00 |
| **APOB** | 10.27 | 8.61 | -1.66 | -3.17 | 0.00 | 0.00 | -1.00 |
| **ALPK3** | 9.18 | 7.51 | -1.67 | -3.18 | 0.00 | 0.00 | -1.00 |
|  | 7.61 | 8.30 | 0.69 | 1.61 | 0.00 | 0.01 | 1.00 |
| **GLUL** | 9.84 | 8.16 | -1.68 | -3.20 | 0.00 | 0.00 | -1.00 |
| **OK/SW-CL.36** | 4.99 | 3.28 | -1.71 | -3.27 | 0.00 | 0.01 | -1.00 |
| **DMRT2** | 8.88 | 7.16 | -1.72 | -3.28 | 0.00 | 0.01 | -1.00 |
| **LOC105376379** | 6.04 | 4.32 | -1.71 | -3.28 | 0.00 | 0.01 | -1.00 |
|  | 3.63 | 4.34 | 0.71 | 1.64 | 0.01 | 0.05 | 1.00 |
|  | 4.58 | 5.61 | 1.03 | 2.04 | 0.00 | 0.01 | 1.00 |
| **TOX3** | 5.68 | 3.95 | -1.73 | -3.32 | 0.02 | 0.08 | -1.00 |
| **EYA1** | 6.63 | 4.89 | -1.74 | -3.35 | 0.00 | 0.00 | -1.00 |
|  | 6.54 | 5.71 | -0.83 | -1.78 | 0.00 | 0.01 | -1.00 |
| **TOX3** | 5.84 | 4.03 | -1.80 | -3.49 | 0.01 | 0.07 | -1.00 |
| **FCN2** | 6.61 | 4.76 | -1.86 | -3.62 | 0.00 | 0.01 | -1.00 |
|  | 4.94 | 5.54 | 0.60 | 1.51 | 0.00 | 0.00 | 1.00 |
|  | 7.65 | 8.62 | 0.97 | 1.96 | 0.00 | 0.01 | 1.00 |
| **COL6A6** | 6.36 | 4.51 | -1.86 | -3.63 | 0.00 | 0.00 | -1.00 |
| **AZGP1** | 11.30 | 9.42 | -1.88 | -3.67 | 0.00 | 0.00 | -1.00 |
| **LOC90768** | 7.53 | 5.65 | -1.88 | -3.68 | 0.00 | 0.00 | -1.00 |
|  | 6.05 | 6.91 | 0.87 | 1.82 | 0.00 | 0.03 | 1.00 |
| **ALDH1L1-AS2** | 7.54 | 5.66 | -1.89 | -3.70 | 0.00 | 0.00 | -1.00 |
| **LDHD** | 8.00 | 6.11 | -1.89 | -3.71 | 0.00 | 0.00 | -1.00 |
| **GLUL** | 8.81 | 6.92 | -1.89 | -3.72 | 0.00 | 0.00 | -1.00 |
| **SPX** | 11.14 | 9.17 | -1.97 | -3.92 | 0.00 | 0.00 | -1.00 |
| **GSDMB** | 8.20 | 6.20 | -2.00 | -4.01 | 0.00 | 0.00 | -1.00 |
| **GPAT3** | 10.43 | 8.40 | -2.03 | -4.09 | 0.00 | 0.00 | -1.00 |
| **PKP2** | 7.81 | 5.76 | -2.05 | -4.14 | 0.00 | 0.01 | -1.00 |
| **RASSF6** | 6.20 | 4.08 | -2.12 | -4.34 | 0.00 | 0.02 | -1.00 |
| **NWD2** | 7.31 | 5.04 | -2.27 | -4.81 | 0.00 | 0.00 | -1.00 |
| **C6** | 10.04 | 7.74 | -2.30 | -4.93 | 0.00 | 0.00 | -1.00 |
| **FAM95A** | 6.42 | 4.09 | -2.33 | -5.02 | 0.00 | 0.00 | -1.00 |
| **COL6A6** | 9.60 | 7.25 | -2.35 | -5.09 | 0.00 | 0.00 | -1.00 |
| **STOX1** | 8.67 | 6.32 | -2.35 | -5.10 | 0.00 | 0.00 | -1.00 |
| **CA3** | 10.47 | 8.02 | -2.45 | -5.46 | 0.00 | 0.00 | -1.00 |
| **RORB** | 6.57 | 3.47 | -3.11 | -8.61 | 0.00 | 0.00 | -1.00 |
| **SPX** | 9.35 | 6.12 | -3.23 | -9.37 | 0.00 | 0.00 | -1.00 |
| **SLC27A2** | 6.94 | 3.56 | -3.38 | -10.42 | 0.00 | 0.00 | -1.00 |
| **RORB** | 8.92 | 5.52 | -3.40 | -10.54 | 0.00 | 0.00 | -1.00 |
| **SLC27A2** | 7.34 | 3.56 | -3.78 | -13.75 | 0.00 | 0.00 | -1.00 |

**Table S5.**
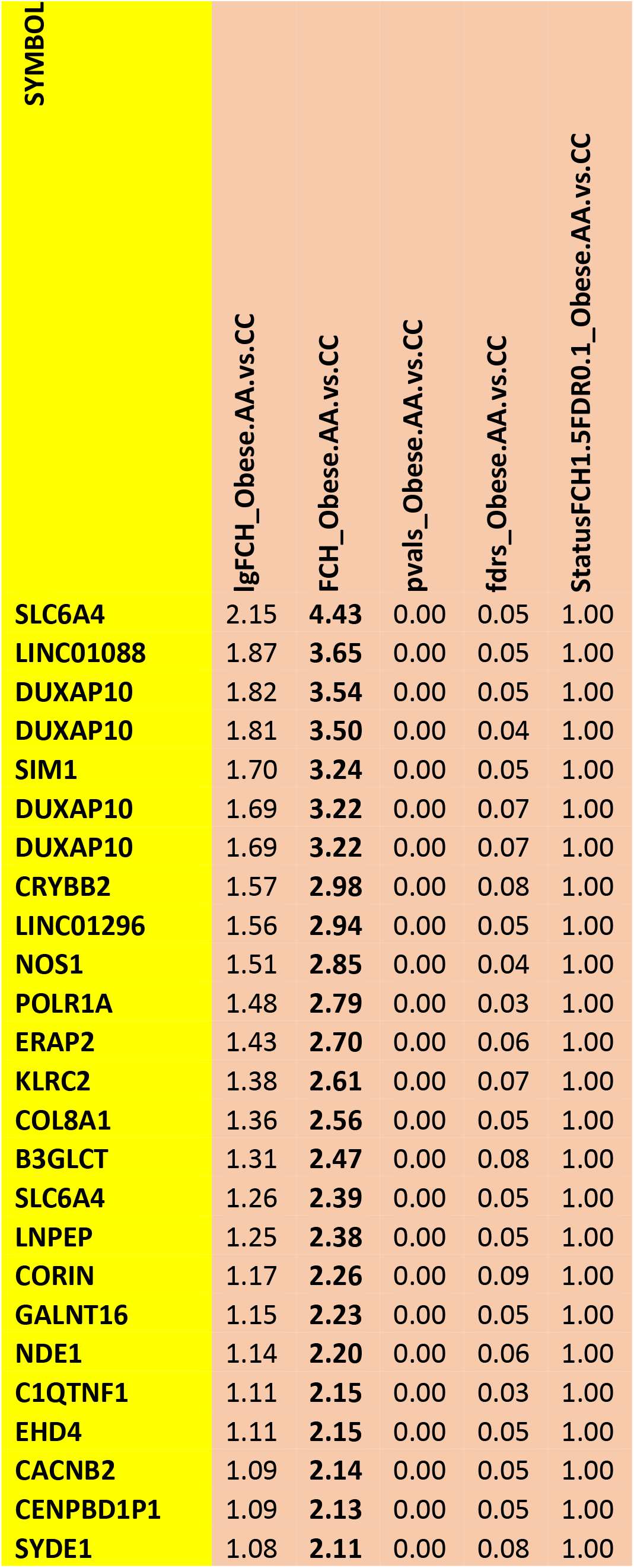
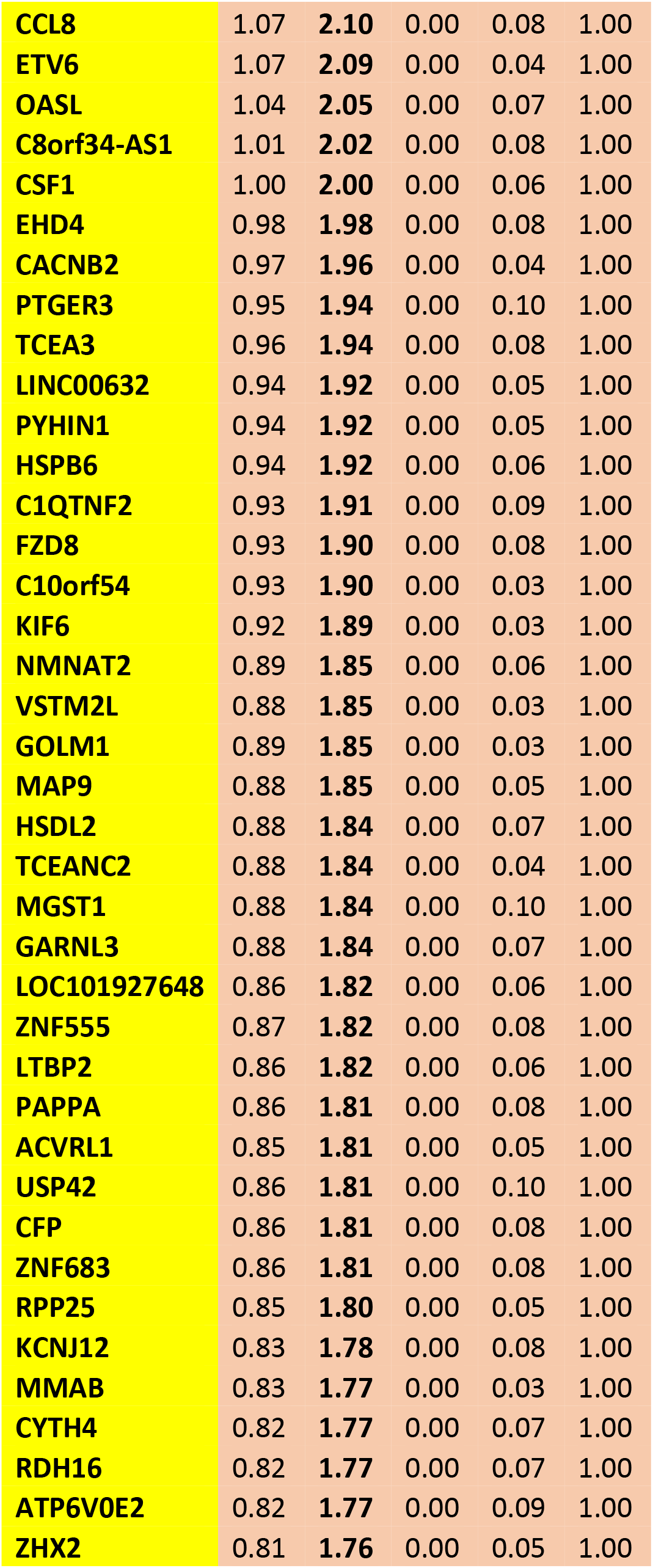
First 70 genes Obese by Race

| SYMBOL |  | lgFCH_Obese.AA.vs.CC | FCH_Obese.AA.vs.CC | pvals_Obese.AA.vs.CC | fdrs_Obese.AA.vs.CC | StatusFCH1.5FDR0.1_Obese.AA.vs.CC |
| --- | --- | --- | --- | --- | --- | --- |
| SLC6A4 |  | 2.15 | **4.43** | 0.00 | 0.05 | 1.00 |
| LINC01088 |  | 1.87 | **3.65** | 0.00 | 0.05 | 1.00 |
| DUXAP10 |  | 1.82 | **3.54** | 0.00 | 0.05 | 1.00 |
| DUXAP10 |  | 1.81 | **3.50** | 0.00 | 0.04 | 1.00 |
| SIM1 |  | 1.70 | **3.24** | 0.00 | 0.05 | 1.00 |
| DUXAP10 |  | 1.69 | **3.22** | 0.00 | 0.07 | 1.00 |
| DUXAP10 |  | 1.69 | **3.22** | 0.00 | 0.07 | 1.00 |
| CRYBB2 |  | 1.57 | **2.98** | 0.00 | 0.08 | 1.00 |
| LINC01296 |  | 1.56 | **2.94** | 0.00 | 0.05 | 1.00 |
| NOS1 |  | 1.51 | **2.85** | 0.00 | 0.04 | 1.00 |
| POLR1A |  | 1.48 | **2.79** | 0.00 | 0.03 | 1.00 |
| ERAP2 |  | 1.43 | **2.70** | 0.00 | 0.06 | 1.00 |
| KLRC2 |  | 1.38 | **2.61** | 0.00 | 0.07 | 1.00 |
| COL8A1 |  | 1.36 | **2.56** | 0.00 | 0.05 | 1.00 |
| B3GLCT |  | 1.31 | **2.47** | 0.00 | 0.08 | 1.00 |
| SLC6A4 |  | 1.26 | **2.39** | 0.00 | 0.05 | 1.00 |
| LNPEP |  | 1.25 | **2.38** | 0.00 | 0.05 | 1.00 |
| CORIN |  | 1.17 | **2.26** | 0.00 | 0.09 | 1.00 |
| GALNT16 |  | 1.15 | **2.23** | 0.00 | 0.05 | 1.00 |
| NDE1 |  | 1.14 | **2.20** | 0.00 | 0.06 | 1.00 |
| C1QTNF1 |  | 1.11 | **2.15** | 0.00 | 0.03 | 1.00 |
| EHD4 |  | 1.11 | **2.15** | 0.00 | 0.05 | 1.00 |
| CACNB2 |  | 1.09 | **2.14** | 0.00 | 0.05 | 1.00 |
| CENPBD1P1 |  | 1.09 | **2.13** | 0.00 | 0.05 | 1.00 |
| SYDE1 |  | 1.08 | **2.11** | 0.00 | 0.08 | 1.00 |

|  |  |  |  |  |  |
| --- | --- | --- | --- | --- | --- |
| **CCL8** | 1.07 | **2.10** | 0.00 | 0.08 | 1.00 |
| **ETV6** | 1.07 | **2.09** | 0.00 | 0.04 | 1.00 |
| **OASL** | 1.04 | **2.05** | 0.00 | 0.07 | 1.00 |
| **C8orf34-AS1** | 1.01 | **2.02** | 0.00 | 0.08 | 1.00 |
| **CSF1** | 1.00 | **2.00** | 0.00 | 0.06 | 1.00 |
| **EHD4** | 0.98 | **1.98** | 0.00 | 0.08 | 1.00 |
| **CACNB2** | 0.97 | **1.96** | 0.00 | 0.04 | 1.00 |
| **PTGER3** | 0.95 | **1.94** | 0.00 | 0.10 | 1.00 |
| **TCEA3** | 0.96 | **1.94** | 0.00 | 0.08 | 1.00 |
| **LINC00632** | 0.94 | **1.92** | 0.00 | 0.05 | 1.00 |
| **PYHIN1** | 0.94 | **1.92** | 0.00 | 0.05 | 1.00 |
| **HSPB6** | 0.94 | **1.92** | 0.00 | 0.06 | 1.00 |
| **C1QTNF2** | 0.93 | **1.91** | 0.00 | 0.09 | 1.00 |
| **FZD8** | 0.93 | **1.90** | 0.00 | 0.08 | 1.00 |
| **C10orf54** | 0.93 | **1.90** | 0.00 | 0.03 | 1.00 |
| **KIF6** | 0.92 | **1.89** | 0.00 | 0.03 | 1.00 |
| **NMNAT2** | 0.89 | **1.85** | 0.00 | 0.06 | 1.00 |
| **VSTM2L** | 0.88 | **1.85** | 0.00 | 0.03 | 1.00 |
| **GOLM1** | 0.89 | **1.85** | 0.00 | 0.03 | 1.00 |
| **MAP9** | 0.88 | **1.85** | 0.00 | 0.05 | 1.00 |
| **HSDL2** | 0.88 | **1.84** | 0.00 | 0.07 | 1.00 |
| **TCEANC2** | 0.88 | **1.84** | 0.00 | 0.04 | 1.00 |
| **MGST1** | 0.88 | **1.84** | 0.00 | 0.10 | 1.00 |
| **GARNL3** | 0.88 | **1.84** | 0.00 | 0.07 | 1.00 |
| **LOC101927648** | 0.86 | **1.82** | 0.00 | 0.06 | 1.00 |
| **ZNF555** | 0.87 | **1.82** | 0.00 | 0.08 | 1.00 |
| **LTBP2** | 0.86 | **1.82** | 0.00 | 0.06 | 1.00 |
| **PAPPA** | 0.86 | **1.81** | 0.00 | 0.08 | 1.00 |
| **ACVRL1** | 0.85 | **1.81** | 0.00 | 0.05 | 1.00 |
| **USP42** | 0.86 | **1.81** | 0.00 | 0.10 | 1.00 |
| **CFP** | 0.86 | **1.81** | 0.00 | 0.08 | 1.00 |
| **ZNF683** | 0.86 | **1.81** | 0.00 | 0.08 | 1.00 |
| **RPP25** | 0.85 | **1.80** | 0.00 | 0.05 | 1.00 |
| **KCNJ12** | 0.83 | **1.78** | 0.00 | 0.08 | 1.00 |
| **MMAB** | 0.83 | **1.77** | 0.00 | 0.03 | 1.00 |
| **CYTH4** | 0.82 | **1.77** | 0.00 | 0.07 | 1.00 |
| **RDH16** | 0.82 | **1.77** | 0.00 | 0.07 | 1.00 |
| **ATP6V0E2** | 0.82 | **1.77** | 0.00 | 0.09 | 1.00 |
| **ZHX2** | 0.81 | **1.76** | 0.00 | 0.05 | 1.00 |

**Table S6.**
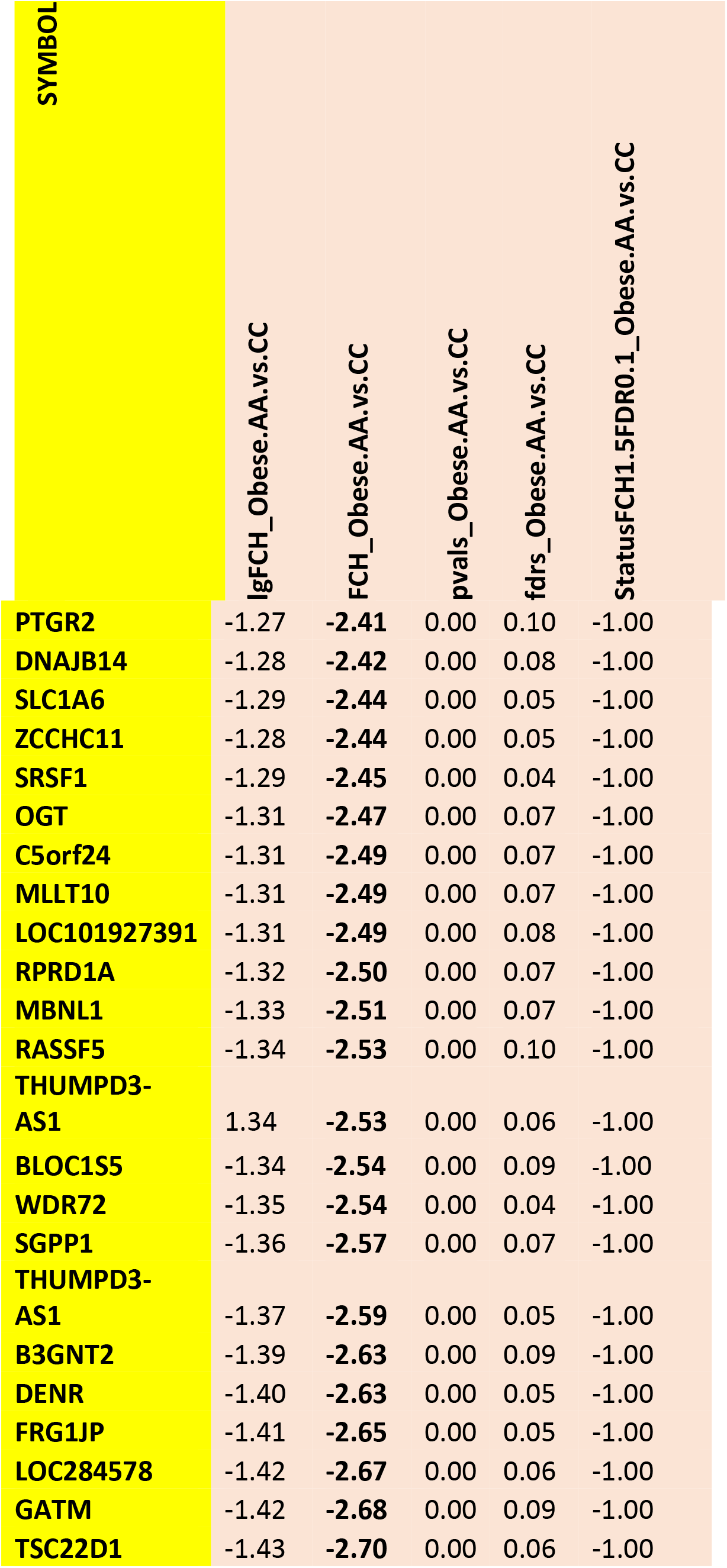
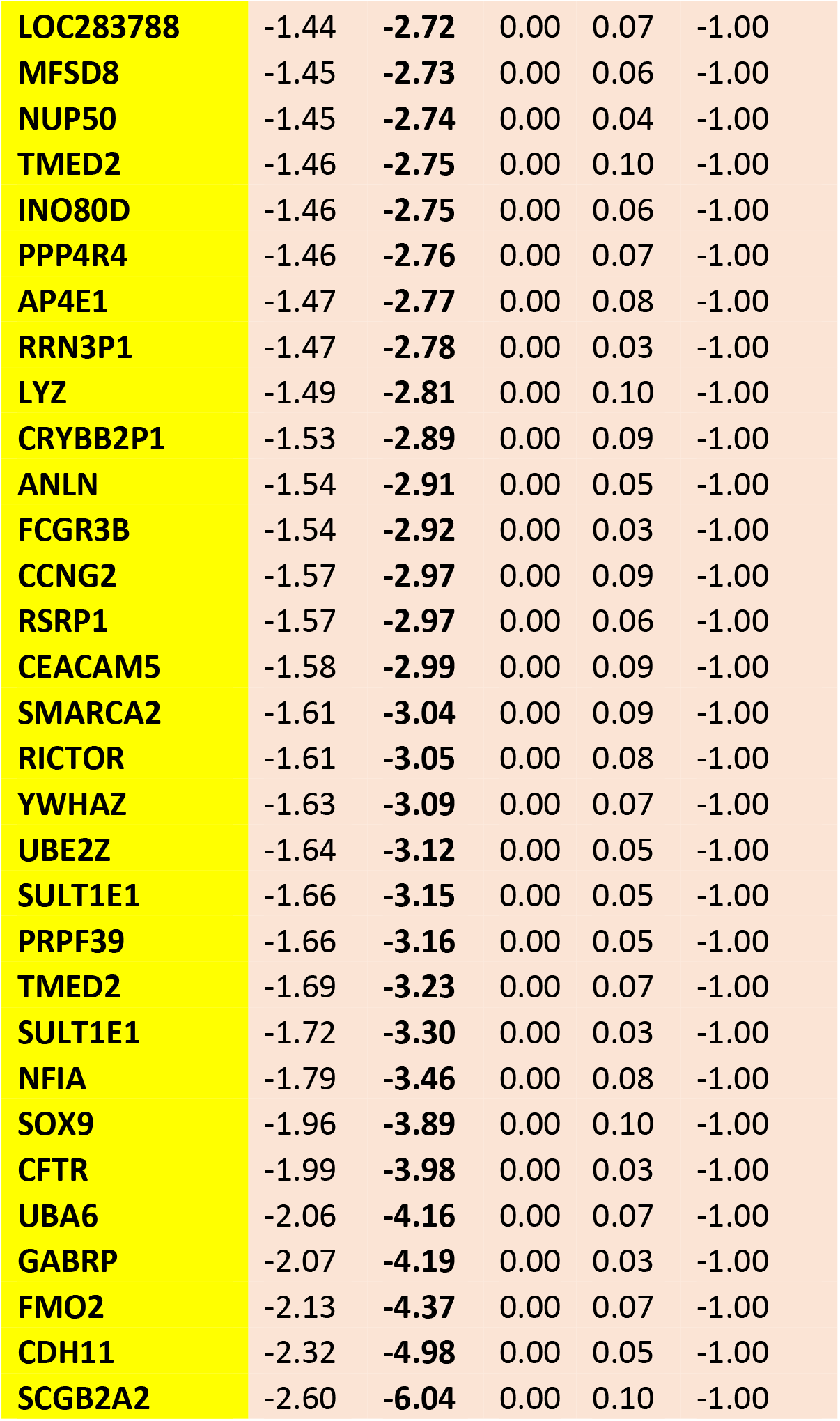
Last 70 genes by Race

**Supplementary Figure S1:**
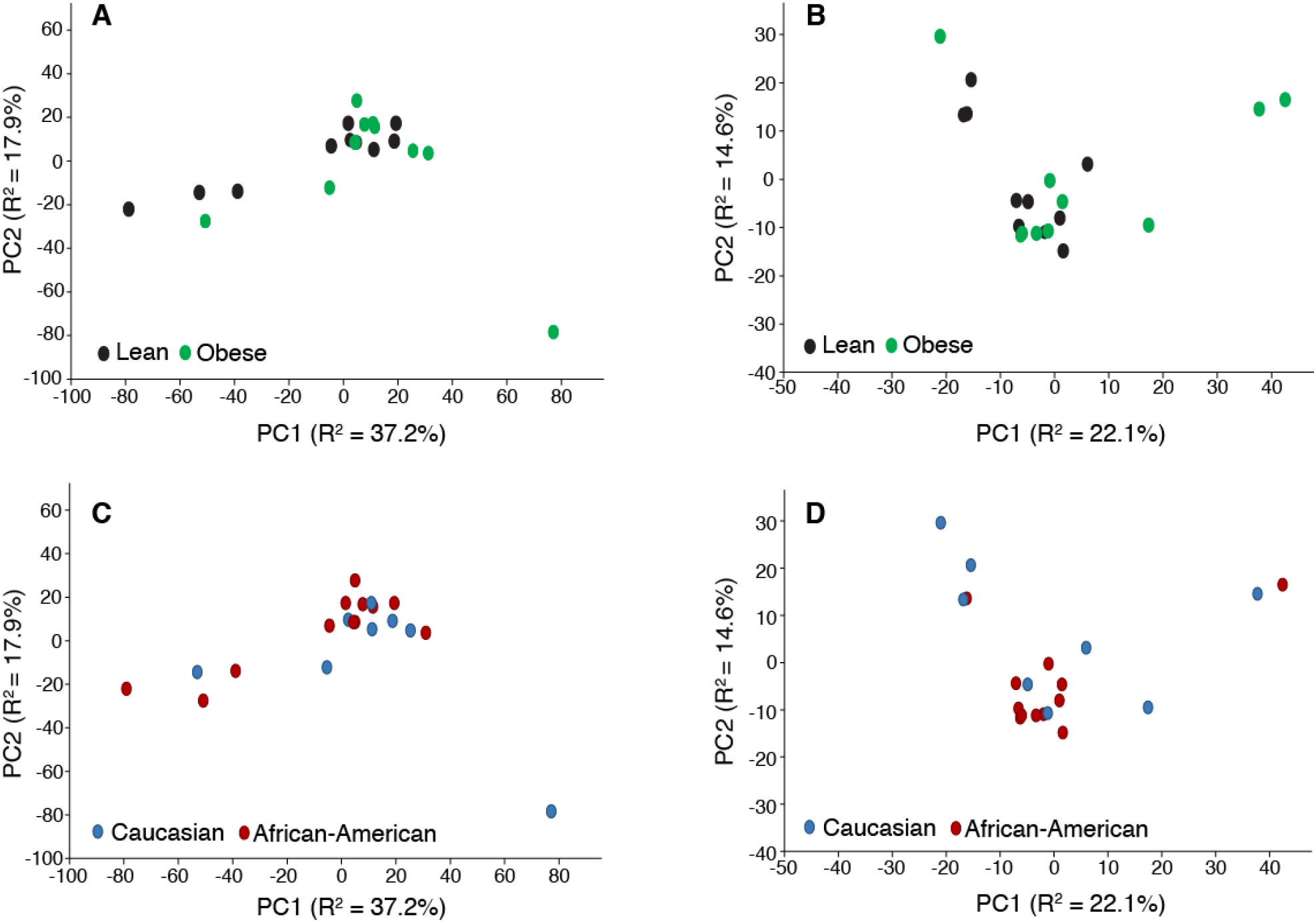
PCA models comparing the metabolic content of swab samples collected from obese and non-obese (A, positive ionization mode; B, negative ionization mode), and Caucasian and African American individuals (C, positive ionization mode; D, negative ionization mode). Metabolic profiles were measured by UPLC-MS.

## Notes

### Competing Interest Statement

The authors have declared no competing interest.

### Clinical Trial

This study does not involve a health-related intervention and therefore does not meet the criteria for registration with ClinicalTrials.gov

### Funding Statement

JMW received research grants from RUCCTS grant#ULTR001866 from the National Center for Advancing Translational Sciences (NCATS), National Institutes of Health (NIH) Clinical and Translational Science Award (CTSA) Program.
JOA received research grants from the Center for Diseases of the Digestive System and Sackler Center for Bionutrition at Rockefeller Universtiy, AHA 17-SFRN33520045 and K08-DK117064 (NIH) at NYU Langone Health.

### Author Declarations

The institutional review board of the Rockefeller University Hospital, and the Advisory Committee for Clinical and Translational Research approved the protocol JWA-0921 after full committee review on 11/30/2016, and thereafter annually.

